# An open, longitudinal resource for mapping interindividual variation in the aging connectome

**DOI:** 10.1101/2025.10.10.25337774

**Authors:** Anna MacKay-Brandt, Yunglin Gazes, Daniel Garcia-Barnett, Lauren Grebe, Olivia Ripley, Kai Xuan Gan, Kristin T. Trautman, Melissa Kramer, Melissa Breland, Russell Tobe, Alexandre Franco, Vilma Gabbay, Michael Milham, Stan Colcombe

**Affiliations:** Brain Aging and Mental Health Laboratory, Clinical Research, Nathan Kline Institute, Orangeburg, NY, USA; Design Acquisition and Neuromodulation Laboratories, Center for Biomedical Imaging and Neuromodulation, Nathan Kline Institute, Orangeburg, NY, USA; Communication Sciences and Disorders, St. John’s University, Queens, NY, USA; Clinical Research, Nathan Kline Institute, Orangeburg, NY, 10962, USA; Child Mind Institute, New York, NY, USA; Department of Psychiatry, New York University Grossman School of Medicine, New York, NY, USA; University of Miami Leonard M. Miller School of Medicine, Miami, FL, USA

## Abstract

Trajectories of age-related neurocognitive decline are not uniform, and are impacted by numerous environmental and physiological factors. Earlier life phases set the stage for later life neurocognitive function, with midlife marking a critical transition characterized by increasing variability in cognitive, affective, and physiological functioning. Despite its importance, this turbulent period remains underrepresented in open neuroimaging and phenotypic data resources. To address this gap, the Nathan Kline Institute - Rockland Sample (NKI-RS) initiative created the ‘Mapping Interindividual Variation in the Aging Connectome’ (MIVAC) substudy—an openly shared, multimodal dataset designed to map brain aging trajectories beginning in midlife and assess the influence of modifiable factors such as cardiorespiratory fitness. This longitudinal investigation includes 348 community-ascertained participants aged 38 to 71 years at baseline. Data collection incorporated deep phenotyping across cognitive, behavioral, medical, and cardiorespiratory fitness domains, along with multimodal neuroimaging (resting-state fMRI, diffusion MRI, morphometric MRI, and arterial spin labeling) and biospecimen collection. The protocol harmonizes with prior NKI-RS substudies while incorporating age-specific considerations for cognitive and neural aging. The full dataset is openly available.

## BACKGROUND

Psychiatric and cognitive disorders are prevalent throughout the lifespan and pose a significant burden on individuals, families, and public health systems. Considerable recent effort has been directed toward understanding disruptions in brain function underlying early-onset psychiatric illness^1–4^ and late-life cognitive decline^3^. However, midlife - despite the emerging recognition that it sets the stage for later life neurocognition^5–7^ - remains a relatively underrepresented period in lifespan neuroscience research^8^. This stage of life, typically defined as the period between the fourth and seventh decades, is characterized by increased variation—both between and within individuals—in cognitive^9,10^, brain^11^, and physiological^12^ functioning. It is also a critical period in which modifiable risk and protective factors may influence later life brain health and cognition^13,14^. Yet, few open-access neuroimaging datasets exist that characterize brain structure, function, and behavior across mid-to-late-life, either cross-sectionally^15^ or longitudinally^16,17^. The Nathan Kline Institute - Rockland Sample (NKI-RS) - Mapping Interindividual Variation in the Aging Connectome’ (MIVAC) substudy was designed to emphasize cardiorespiratory fitness and physiological function across multiple timepoints to understand the impact of natural variation of key modifiable factors on brain and cognitive function in community samples.

The current dataset was designed to complement the previously described pediatric longitudinal protocol of the NKI-RS^4^, which emphasized brain development and the emergence of psychiatric illness from ages 6 to 17. This substudy extends the community-ascertained, transdiagnostic, and deeply phenotyped framework of the NKI-RS program^18^ for a longitudinal characterization of interindividual variation across midlife and older adulthood (ages 38–71).

The MIVAC substudy supports efforts to map normative and atypical brain aging trajectories and to evaluate modifiable determinants of cognitive and neural health. Specifically, this study incorporates cardiorespiratory fitness assessment, medical and behavioral phenotyping, and high-resolution multimodal imaging to investigate risk and protective factors that affect brain health and cognition.

To achieve these goals, we implemented a multi-cohort longitudinal design, prospectively harmonized our core protocol with prior NKI-RS substudies, and maintained the NKI-RS commitment to open science and data sharing. Imaging protocols include multiband functional and high-angular resolution diffusion magnetic resonance imaging (MRI), arterial spin labeling, and structural morphometry^4^. Comprehensive phenotyping includes psychiatric diagnostic interviews, cognitive and behavioral assessments, medical history, laboratory tests, physical measures, and the addition of gold-standard cardiorespiratory fitness assessment (VO_2_max). Protocol harmonization across prior lifespan cross-sectional and pediatric longitudinal NKI-RS substudies allows for unique, deeply-phenotyped, lifespan neuroimaging analyses. This data descriptor outlines the rationale, design, implementation, and available data for the NKI-RS MIVAC cohort and provides guidance for researchers interested in the use of these data for the study of brain aging from midlife to older adulthood.

## METHODS

### Recruitment and Retention Strategy

The target enrollment was 434, with 400 participants recruited between January 2016 and April 2018. The last follow-up assessment completed before the coronavirus pandemic-related shutdown was in March 2020. Recruitment followed the strategies established across all NKI-RS substudies to approximate a community-representative enrollment for the local region. Detailed recruitment descriptions common to all NKI-RS studies can be found in^18^ and ^4^.

In addition to program-wide approaches, the MIVAC substudy included unique recruitment, retention, and feedback strategies. Study investigators with expertise in healthy aging hosted community talks at NKI and in local community spaces. Limited clinically-relevant feedback was provided to participants upon request to support the individual health goals of our participants when shared with their healthcare providers (See^4^ for more detail). However, due to concerns about sampling bias, feedback was not highlighted as part of MIVAC study recruitment and potential participants who stated concern about memory impairment were referred for a free memory evaluation through the NKI Geriatric Psychiatry Research Program and not enrolled.

Building upon lessons learned from three preceding NKI-RS substudies^4,18,19^, procedures adopted or implemented to promote retention included:

- Birthday cards sent to participants to help maintain a positive connection and track address changes.
- Open house events and “coffee talks” hosted by the study investigators and designed to create a dialogue to translate preliminary findings, report on study progress, and answer any general questions about healthy aging raised by the participant community.
- Newsletters emailed to participants.
- Written and verbal participant satisfaction collected at each study visit.
- Prior to MIVAC data collection, the core protocol across all prior substudies was reviewed by the NKI-RS leadership team and reduced to increase efficiency and decrease participant burden.

Of the 400 participants enrolled, 348 (87%) completed a baseline visit. Among those who completed baseline visits, 5 (1.4%) were identified to have an MRI contraindication and were excluded. Of the remaining participants, 264 (77%) completed a first annual follow-up. A second annual follow-up was completed by 219 (64%) participants. The coronavirus pandemic interrupted the third annual follow-up period, with 106 (31%) completed, 16 (5%) due for assessment but lost to follow-up, and 97 (28%) scheduled but administratively canceled for health safety reasons related to pandemic precautions. Of note, 285 (82%) study participants completed an online survey during the first three months of the coronavirus pandemic-related shutdown (May 2020 - August 2020) reporting on the impact of the public health mandated shutdown on their daily life activities, physical and psychological health. See Figure 1 for visualizations of age by visit and a Strengthening the Reporting of Observational Studies in Epidemiology (STROBE) reporting diagram for observational studies^20^.

**Figure 1.**
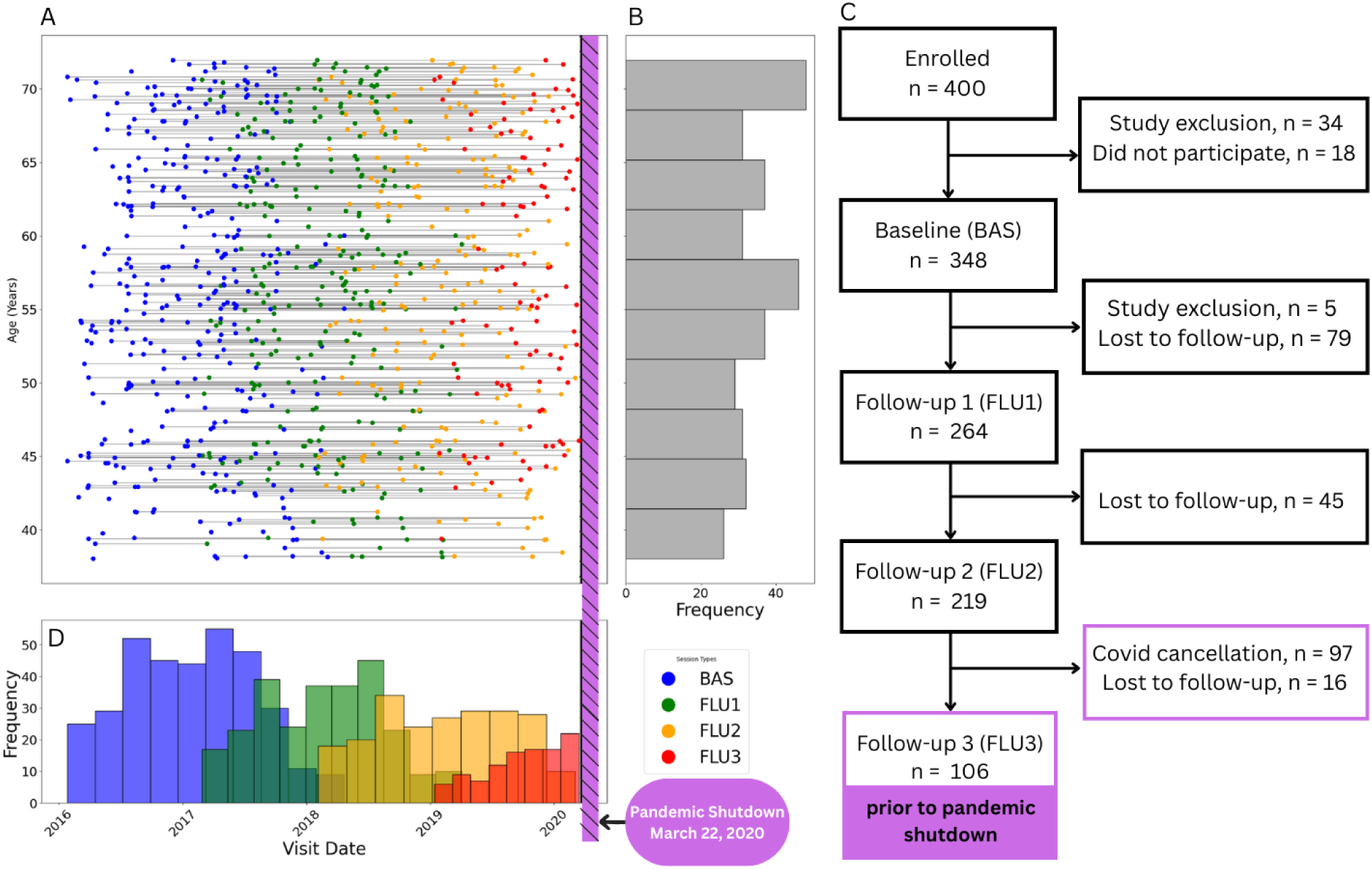
Overview of participant age distribution and retention. A. Timeline for each participant’s visits plotted against their age. Each visit is color-coded for visit number. B. Histogram of participant age. C. STROBE reporting diagram for observational studies. D. Histogram of each visit. Hatched purple bar indicates the date of pandemic shutdown on March 22, 2020. 285 out of 348 participants from BAS (82%) participated in the COVID survey during the pandemic shutdown. Participant Procedures.

All NKI-RS substudies followed the same procedures, with staff trained across all contemporaneous studies and supervised by the NKI-RS leadership team. Common procedures are detailed in ^4,18^ and available on the NKI-RS website (https://rocklandsample.org/for-researchers). The MIVAC substudy specific procedures are described in detail below.

### Screening

Potential participants completed a pre-screening phone interview with an intake coordinator. The screening interview assessed eligibility across all NKI-RS substudies according to common and study specific inclusion criteria (See Figure 2). Individuals meeting study criteria were invited to participate.

**Figure 2.**
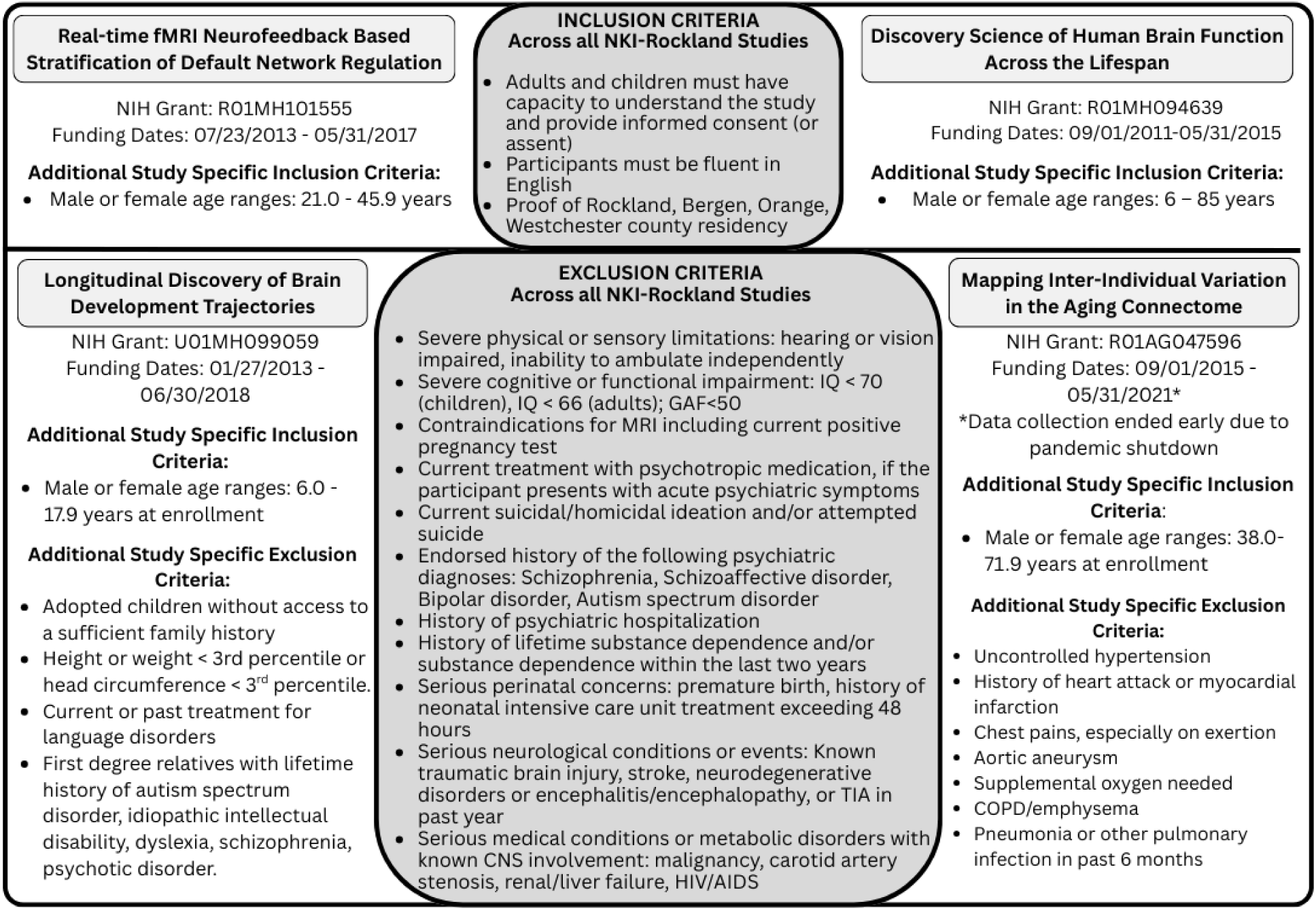
Inclusion and exclusion criteria for the aggregate NKI-RS cohort and each substudy. Shared selection criteria among the four studies are presented in the center boxes. Study specific criteria are listed under each NKI-RS substudy.

### IRB approval

This study was approved by the Nathan Kline Institute for Psychiatric Research Institutional Review Board. Prior to conducting the research, written informed consent was obtained from the participants.

### Experimental Design

The MIVAC study design included 9 visits across four years. In the first year, participants attended a brief consent visit, a baseline full-day characterization visit, and a separate one-hour cardiorespiratory fitness assessment. Each annual follow-up included separate characterization and cardiorespiratory fitness assessment visits. Visits were conducted on consecutive days, when possible, with no more than 2 weeks separation if necessary. Adjustments to the schedule were made to complete assessments on an additional visit when necessary to decrease participant burden. Datasets include a variable that codes for interval time between each assessment (i.e. “day lag”) to allow researchers greater precision when considering analyses within a data collection epoch (e.g. baseline visit). All assessments are listed in Table 1. An example schedule is provided in Table 2; See also https://rocklandsample.org/mivac-full-end-user-protocol-2 for full details.

**Table 1.** Assessments by visit.

| Assessments | Baseline | Follow-Up | Assessments | Baseline | Follow-Up |
| --- | --- | --- | --- | --- | --- |
| **General** |  |  | **Imaging** |  |  |
| Brain Injury Screening Questionnaire (BISQ) | ✓ | ✓ | MRI | ✓ | ✓ |
| Consent | ✓ | ✓ | MRI-Questionnaire | ✓ | ✓ |
| Demographics Questionnaire | ✓ | ✓ | **Diagnostic** |  |  |
| Demographics Supplement | ✓ | ✓ | Adult ADHD Clinical Diagnostic Scale (ACDS) | ✓ |  |
| Family History Questionnaire | ✓ |  | Global Outcome Scale (GOS; 65+) | ✓ | ✓ |
| Hollingshead Socioeconomic Status (SES) | ✓ |  | Structured Clinical Interview for DSM Disorders (SCID) | ✓ | ✓ |
| Medical History Questionnaire | ✓ | ✓ | **Behavioral Tasks** |  |  |
| Medications/Medical Conditions | ✓ | ✓ | Rapid Visual Information Processing (RVIP) | ✓ | ✓ |
| Satisfaction Survey | ✓ | ✓ | **Behavior and Phenotypic Characterization Questionnaires** |  |  |
| Social Networking Questionnaire | ✓ | ✓ | Adult Self Report / Older Adult Self Report Adaptive Functioning (ASR-AF/OASR-AF) | ✓ | ✓ |
| Zipcodes | ✓ | ✓ | Adult Self Report / Older Adult Self Report (ASR/OASR) | ✓ | ✓ |
| **Physical** |  |  | Adult Temperament Questionnaire (ATQ) | ✓ | ✓ |
| 6-Minute Bike Test | ✓ |  | Beck Depression Inventory (BDI - II) | ✓ | ✓ |
| Actigraphy | ✓ |  | Cognitive Failures Questionnaire (CFQ) | ✓ | ✓ |
| Blood Collection | ✓ | ✓ | Conners Adult ADHD Rating Scale – Self Report, Short Version (CAARS) | ✓ | ✓ |
| Cambridge-Hopkins Restless Legs Syndrome (CHRLS) | ✓ | ✓ | Domain-Specific Risk-Taking Scale (DOSPERT) | ✓ | ✓ |
| Color Vision | ✓ |  | Eating Disorder Examination Questionnaire (EDEQ) | ✓ | ✓ |
| Edinburgh Handedness Questionnaire (EHQ) | ✓ |  | Emotional Regulation Questionnaire (ERQ) |  | ✓ |
| Grip Strength | ✓ | ✓ | Inventory of Callous-Unemotional Traits (ICU) | ✓ |  |
| Grooved Pegboard | ✓ | ✓ | Interpersonal Reactivity Index (IRI) | ✓ | ✓ |
| International Physical Activity Questionnaire (IPAQ) | ✓ | ✓ | NEO Five Factor Inventory (NEO-FFI-3) | ✓ | ✓ |
| Pittsburgh Sleep Quality Index (PSQI) | ✓ | ✓ | Physical Activity Scale for the Elderly (PASE) | ✓ | ✓ |
| VO2 Submax Testing* | ✓ | ✓ | 21-Item Peters et al. Delusions Inventory (PDI-21) | ✓ | ✓ |
| Walking While Talking (WWT)* | ✓ | ✓ | PhenX Sexual History | ✓ | ✓ |
| Weight/Height, Vital Signs, Hip/Waist Measurement | ✓ | ✓ | Penn State Worry Questionnaire (PSWQ) |  | ✓ |
| **Cognitive** |  |  | Perseverative Thinking Questionnaire (PTQ) |  | ✓ |
| Attention Network Task (ANT) | ✓ | ✓ | Ruminative Response Scale (RRS) | ✓ | ✓ |
| Digit Span | ✓ | ✓ | Sex Role Identity Scale | ✓ |  |
| Delis-Kaplan Executive Function System (DKEFS) - Color-Word | ✓ | ✓ | Sexual Orientation Scale | ✓ |  |
| DKEFS - Design fluency | ✓ | ✓ | State Trait Anxiety Inventory (STAI) | ✓ | ✓ |
| DKEFS - TOWRE | ✓ | ✓ | Three-Factor Eating (TFEQ) | ✓ | ✓ |
| DKEFS - Trails | ✓ | ✓ | Trauma Symptom Checklist for Adults (TSC-40) | ✓ | ✓ |
| DKEFS - Verbal Fluency | ✓ | ✓ | University of California at Los Angeles Posttraumatic Stress Disorder Reaction Index (UCLA-PTSD) | ✓ | ✓ |
| Montreal Cognitive Assessment (MoCA) | ✓ | ✓ | UPPS-P Impulsive Behavior Scale (UPPS) | ✓ |  |
| Penn Computerized Neurocognitive Battery (CNB) | ✓ | ✓ | **Substance Use and Addiction Measures** |  |  |
| Rey Auditory Verbal Learning Test (RAVLT) | ✓ | ✓ | Comprehensive Addiction Severity Index for Adolescents (CASI-A) | ✓ | ✓ |
| Wechsler Abbreviated Scale of Intelligence (WASI-II) | ✓ | ✓ | Fagerstrom Test for Nicotine Dependence (FTND) | ✓ | ✓ |
| Wechsler Individual Achievement Test – Second Edition Abbreviated (WIAT-IIA) | ✓ |  | National Institute on Drug Abuse Questionnaire (NIDA) | ✓ | ✓ |
|  |  |  | Urine Drug Test | ✓ | ✓ |

\* Assessments unique to MIVAC Study

**Table 2.** Sample Baseline Characterization Schedule. Annual follow-up appointments maintained the same visit schedule, see. Table 1 **for follow-up assessment details.**

| Duration (mins) | Activities |
| --- | --- |
| First Visit - Consent, Brief Cognitive and Health Screening (1 hour) |  |
| 20 | Consent |
| 20 | Cognitive Screening - MoCA |
| 10 | Health Screening - Morphometrics |
| 10 | Set up at-home surveys |
| Second Visit - Main Protocol (Full day) |  |
| 10 | Phlebotomy |
| 15 | Breakfast |
| 90 | Experimental cognitive tasks |
| 10 | Break |
| 90 | Neuroimaging |
| 30 | Lunch |
| 90 | Neuropsychological assessment |
| 60 | Psychiatric diagnostic interview |
| 30 | Psychosocial/health surveys |
| 15 | 6-min bike assessment |
| Third Visit - Fitness assessment (1 hour) |  |
| 15 | Walking While Talking procedure |
| 30 | VO<sub>2</sub> Submaximal Cardiorespiratory Fitness |

### Clinical Assessments

Detailed procedures for clinical assessments common across the NKI-RS substudies can be found in^4^ and https://rocklandsample.org/mivac-full-end-user-protocol-2. Please note that some self-report assessments developed for specific age cohorts were extended to younger or older ages across the NKI-RS aggregate sample (e.g., Montreal Cognitive Assessment: MoCA, Comprehensive Addiction Severity Index for Adolescents: CASI-A), while age-specific versions of measures were also chosen and matched to the participant age (e.g., Achenbach System: Adult Self Report / Older Adult Self Report: ASR, OASR; Geritaric Depression Scale: GDS). The procedure was to assign sets of surveys to participants based on age that balanced breadth of coverage for the aggregate NKI-RS lifespan sample while also recognizing the importance of age-specific framing for some measures. Note that some items of the ASR and OASR overlap, allowing for item-level coverage across middle to older adulthood. See Supplementary Tables 1 and 2 for details.

Unique to the MIVAC substudy was the characterization of global cognitive function using the MoCA^21^. Research technicians, under the supervision of the study neuropsychologist, administered the MoCA at baseline and each annual follow-up visit. Administration and scoring followed the standardized protocol. All scores below 26 were sent to the neuropsychologist for review. Scores were compared with all other available study data and any contextual information.

### MRI

All NKI-RS substudies shared the same core neuroimaging protocol, and collection was performed using the same 3.0T Siemens TIM Trio scanner at the Nathan Kline Institute with a 32-channel head coil for all acquisitions. During each imaging session, participants underwent a comprehensive protocol consisting of two high resolution morphometric sequences (T1-weighted and T2-SPACE), one diffusion MRI sequence, six functional MRI runs, one arterial spin labeling (ASL), and a T2-FLAIR sequence. Imaging parameters are presented in Table 3. See also^4^ for a detailed description of this core NKI-RS protocol. Full imaging protocols can be downloaded here: https://rocklandsample.org/for-researchers-rockland-sample-i-2011-2022-neuroimaging-protocol

**Table 3.** Parameters for MR Sequences. MPRAGE = Magnetization Prepared Rapid Gradient Echo; SPACE = Sampling Perfection, Application optimized Contrast; FLAIR = Fluid-Attenuated Inversion Recovery; dMRI = diffusion MRI; R-fMRI = Resting state functional MRI; mb = Multiband; sb = Singleband; pCASL = Pseudo-Continuous Arterial Spin Labeling; FOV = Field of View; TR = Repetition time; TE = Echo time; TI = Inversion time.

| Scan Type | Time Acquisition (min:sec) | Slices | % FOV Phase | Resolution (mm) | matrix | TR (ms) | TE (ms) | Flip Angle (°) | Multi Band Accel | Phase Partial Fourier | Notes |
| --- | --- | --- | --- | --- | --- | --- | --- | --- | --- | --- | --- |
| T1-weighted (MPRAGE) | 4:18 | 176 | 96% | 1 × 1 × 1 | 256 × 256 | 1900 | 2.52 | 9 | N/A | off | TI (ms) = 900 |
| T2 FLAIR | 3:02 | 44 | 75% | 1 × 1 × 3 | 256 × 192 | 900 | 106 | 180 | N/A | off | TI (ms) = 2500 |
| dMRI | 5:43 | 64 | 84.90% | 2 × 2 × 2 | 106 × 90 | 2400 | 87 | 90 | 4 | 6/8 | 128 directions; b = 1500 s/mm<sup>2</sup>; 9 b0 images |
| R-fMRI (mb-645: Rest-645) | 9:46 | 40 | 100% | 3 × 3 × 3 | 74 × 74 | 645 | 30 | 60 | 4 | off |  |
| R-fMRI (mb-1400: Rest-1400) | 9:45 | 64 | 100% | 2 × 2 × 2 | 112 × 122 | 1400 | 30 | 65 | 4 | 6/8 |  |
| R-fMRI (sb: Rest-CAP) | 5:05 | 38 | 100% | 3 × 3 × 3 | 72 × 72 | 2500 | 30 | 80 | sb | off |  |
| Visual Checkerboard Stimulation (CB-645) | 2:41 | 40 | 100% | 3 × 3 × 3 | 74 × 74 | 645 | 30 | 60 | 4 | off |  |
| Visual Checkerboard Stimulation (CB-1400) | 2:27 | 64 | 100% | 2 × 2 × 2 | 112 × 112 | 1400 | 30 | 64 | 4 | 6/8 |  |
| Breath Holding (BH-1400) | 4:30 | 64 | 100% | 2 × 2 × 2 | 112 × 112 | 1400 | 30 | 65 | 4 | 6/8 |  |
| pCASL | 5:15 | 24 | 100% | 3.4 × 3.4 × 4.2 | 64 × 64 | 3800 | 17 | 90 | N/A | 7/8 | Label offset = 80 mm, post label delay = 1000 ms |
| T2 SPACE | 3:52 | 176 | 100% | 1 × 1 × 1 | 256 × 256 | 3200 | 305 | varies | N/A | off |  |

### Physical Health Assessments

All NKI-RS substudies followed the same procedures for assessing height, weight, waist/hip circumference, heart rate, blood pressure, grip strength, vision, and activity levels/sleep. Routine venipuncture assays were acquired at each visit as part of the physical-health protocol for laboratory analysis and to contribute to the biospecimen bank at the NIMH Genetics Repository. See^4^ for a detailed description of those procedures.

Unique to the MIVAC substudy, participant cardiorespiratory fitness and walking speed were assessed at baseline and each follow-up visit. Trained study research technicians, under the supervision of MPI Colcombe, administered gold standard submaximal VO_2_max assessments using a recumbent stationary bike connected to the Parvo Medics True One 2400 metabolic measurement system^22^. During the testing session, participants wore a fitted silicon mask attached to the metabolic measurement system, quantifying the relative concentrations of oxygen and CO_2_ in their exhaled breath gasses, as well as a sensor to measure heart rate.

Participants pedaled with the aim of reaching at least 90% of their age-predicted maximum heart rate (220-age) and achieving a respiratory exchange ratio (RER; CO_2_ to O_2_ ratio) of 1.02 or higher. During administration, medical support was provided by on-site clinicians. Cardiorespiratory fitness data include data quality criteria.

Walking speed alone and under cognitive load was assessed as a low tech but clinically scalable counterpart to the gold-standard cardiorespiratory fitness assessment and has been shown to predict falls, disability, and cognitive decline^23^. Using a protocol developed by researchers at the Albert Einstein College of Medicine^24^, participants were instructed to walk at their normal pace along a 20-foot distance marked off with tape, recite alternate letters of the alphabet, and walk while reciting alternate letters of the alphabet. Two trials were completed for each condition. Walking time, total correct letters, and total errors were recorded by a trained research technician for each trial across single and dual task conditions. See https://rocklandsample.org/mivac-full-end-user-protocol-2 for full details of both physical assessments.

### Data Distribution and Use

#### Maintenance of End User Inquiry Mechanism

As of July 2025, 349 collaborative research sites have obtained full access to the enhanced NKI-RS dataset by signing a Data Use Agreement (DUA). An inquiry response mechanism to ensure “End Users” appropriately understand and access data was developed and retained after study completion. End users submit inquiries to for support with data access, data quality, or documentation; to provide feedback if a technical error is identified; and to document publications using the data resource. An inquiry mechanism is critical for effective use of these data resources. Accordingly, an End User Response Panel—composed of NKI-RS program investigators with expertise in phenotyping, clinical characterization, database management, and imaging—was developed. NKI-RS research staff monitor email at least weekly and forward questions to the Panel. Since implementation, the Panel has received and jointly responded to 598 unique inquiry threads (2017: 83; 2018: 62; 2019: 41; 2020: 60; 2021: 81; 2022: 108; 2023: 58; 2024: 56; 2025: 49).

## DATA RECORDS

### Dataset Deposition

All NKI-RS data files, including the MIVAC substudy, are accessible via the International Neuroimaging Data-Sharing Initiative (INDI) under the title “Enhanced Nathan Kline Institute - Rockland Sample (NKI-RS)”^25^.

### Data Privacy

Across all NKI-RS studies, participants provided informed consent for their data to be shared via IRB-approved protocols^4^. Informed consent for sharing via INDI was obtained prior to participation in the study. Given the critical priority of protection of participant privacy, no protected health information is ever released via the INDI. For imaging data, face information was removed from anatomical scans, and protected/identifying information was removed from all image headers and databases. Random 7-digit INDI identifiers were assigned to all datasets. Recognizing the risks associated with sharing phenotypic data, which is high dimensional and personal in nature, the NKI-RS utilizes two release versions:

### NKI-Rockland Lite

This release version contains only de-identified imaging data and limited phenotyping (e.g. age, sex, handedness). The NKI-Rockland Lite is immediately available without DUA requirements. To access the NKI-RS Lite release, please visit the study website (rocklandsample.org).

### NKI-Rockland Full Phenotypic Release

This release contains the full high-dimensional phenotypic protocol common across all NKI-RS substudies. Users of the NKI-Rockland Full Phenotypic Release complete a DUA and obtain appropriate institutional approval prior to accessing data (https://rocklandsample.org/phenotypic-data). The entire phenotypic dataset is available to approved users via the COINS database system^26^ (used for data collection) and the Longitudinal Online Research and Imaging System^27^ (LORIS) database (maintained by NKI-RS program staff as a curated resource).

### Data acknowledgement

A guiding principle for data-sharing across all NKI-Rockland Sample studies is to provide researchers with the greatest flexibility to carry out scientific inquiries. In this regard:

1. We do not require authorship on any manuscripts generated using NKI-Rockland datasets; citation as a data-source is sufficient.
2. We do not require review of manuscripts generated using NKI-Rockland datasets; we do strongly encourage careful documentation of selection criteria applied to the NKI-RS in choosing datasets for analysis.
3. We do not require specification of data analyses prior to data use.

### Distribution for use

The primary website for all NKI-Rockland Initiative studies, including MIVAC, is located at: https://rocklandsample.org. Phenotypic data may be accessed through either an NKI-RS dedicated instance of LORIS located at https://data.rocklandsample.rfmh.org/ and supported by the NKI-RS research program staff or through the COINS Data Exchange (https://coins.trendscenter.org/). Except for the limited phenotypic release, which is publicly available with the imaging, NKI-RS phenotypic data are protected by a DUA. Investigators must complete a DUA (found at https://rocklandsample.org/phenotypic-data) and have it approved by an authorized institutional official before receiving access. The intent of the DUA is to ensure that data users (1) agree to protect participant confidentiality when handling the high dimensional NKI-RS phenotypic data and (2) agree to take the necessary measures to prevent breaches of privacy. The DUA does not place any constraints on the range of analyses that can be carried out using shared data, nor does it include requirements for review or co-authorship by the originators of the NKI-RS.

### Imaging data

All imaging data can be accessed through the Rockland Sample portal (https://rocklandsample.org/accessing-the-neuroimaging-data-releases). This website provides instructions for users to directly download data from an Amazon Simple Storage Service (S3) bucket. These data are organized in the Brain Imaging Data Structure (BIDS) format. We provide two scripts to perform batch download of these imaging files, in Bash script and Python. The Python script has the capability to perform some basic filtering prior to downloading data: for example, sex = female, age >=45, session = BAS1, image type = anat. Instructions are also provided to download individual files using open-source S3 bucket browsers.

### Partial and missing data

Some participants were not able to successfully complete all components of the NKI-RS protocol due to a variety of factors (e.g.,unexpected discomfort in the scanner, family emergency interrupting a testing day). To prevent data loss due to participant discomfort during an MRI session, we included a mock MRI scanner experience during the first visit, when possible. Overall, we attempted to collect as much data as possible within the allotted data collection intervals and logged data losses as they occurred.

### Data license

All NKI-Rockland Sample data are distributed using the Creative Commons-Attribution-Noncommercial license, which is described at: https://creativecommons.org/licenses/by-nc/4.0/legalcode. For the high-dimensional phenotypic data, all terms specified by the DUA must be complied with.

## TECHNICAL VALIDATION

### Sample Composition

#### Geographic Distribution

The Nathan Kline Institute (NKI) is located 25 miles north of New York City in Rockland County. From 2013 through 2019, the NKI-RS initiative enrolled residents of Rockland and neighboring counties (Bergen County, NJ; Orange County, NY; and Westchester County, NY). A previously published report compared the aggregate NKI-RS dataset (n = 1,610) with the demography of Rockland County, New York State, and the United States based on census data and demonstrated similar distributions across the research sample, local, regional, and national demography in most domains^4^. Here we present the MIVAC cohort compared with US census data for additional comparison (Table 4). In the current study, similar to reporting on the aggregate dataset, individuals who lived closer to NKI were more likely to enroll. See Supplementary Figure 1.

**Table 4.** Demographics of the NKI-RS MIVAC sub-study (enrollment years 2016-2019) and United States Census Data.

|  | **NKI-Rockland Sample**<br>**Mapping Inter-**<br>**Individual Variation in**<br>**the Aging**<br>**Connectome** | **USA** |
| --- | --- | --- |
| **Population (estimate)** | 348 | 309,529,237 |
| **Persons 65 years old and over** | 22.70% | 13% |
| **Female persons** | 71.30% | 51% |
| **White** | 83.30% | 79.60% |
| **Black or African American** | 10.10% | 13% |
| **American Indian/Alaska Native** | 0.60% | 1% |
| **Asian** | 4.60% | 5% |
| **Native Hawaiian or Other Pacific Islander** | 0.30% | 0% |
| **Two or more races** | N/A | 2% |
| **Hispanic or Latinx** | 8.00% | 16% |
| **Foreign born** | N/A | 5% |
| **Bilingual at home** | 27.16% | 18% |
| **High school graduates** | 98.80% | 80% |
| **Bachelor's degree or higher** | 69.30% | 24% |
| **Persons per household** | 2.9 | 2.51 |
| **Median household income** | \$75,000 - \$99,900 | \$52,029 |
| **Per capita money income** | N/A | \$21,587 |
| **Persons below poverty level** | N/A | 12.20% |

#### Age, Sex, and Clinical Characterization

Age distributions (Figure 1b) were relatively even across the middle to older adult age range, despite known challenges recruiting working age adults into time intensive research protocols. The contemporaneous recruitment effort for the pediatric longitudinal substudy allowed parents to learn about the MIVAC study and gain direct experience with the research staff, facilitating enrollment across both substudies. Of the 348 participants who completed baseline assessments, 248 (71%) were female. Midlife and older adult participation among men tends to lag behind female participation in studies of healthy aging. ^28^ Cross enrollment with the pediatric substudy may have further contributed to this gender gap, as mothers were often the accompanying guardian.

Clinical characterization of global cognition was conducted as a brief screening using the MoCA. Across 808 total assessments, there were 543 MoCA scores above 25 (67%), 259 scores between 25 and 18 (32%), and 6 scores below 18 (.7%). There was no supporting evidence for dementia among the 4 participants with scores below 18. According to standard cutoff criteria,^21^ 223 (65%) participants were characterized as cognitively normal at baseline (MoCA > 25), and 120 (35%) participants scored in the range suggestive of mild or moderate cognitive impairment at baseline. No study participants scored in the severe range (0-9). Of the 257 participants with baseline and at least one follow-up visit, change of classification was observed in 27% of participants, 28 (11%) fell from “normal” to “mild,” 39 (15%) rose from “mild” to “normal” classification, and 3 (.01%) rose from “moderate” to “mild.” All 4 participants who scored in the “moderate” range (10-17) scored higher at their final follow-up visit. See Figure 3A for an illustration of classification change from baseline to final follow-up visit.

**Figure 3.**
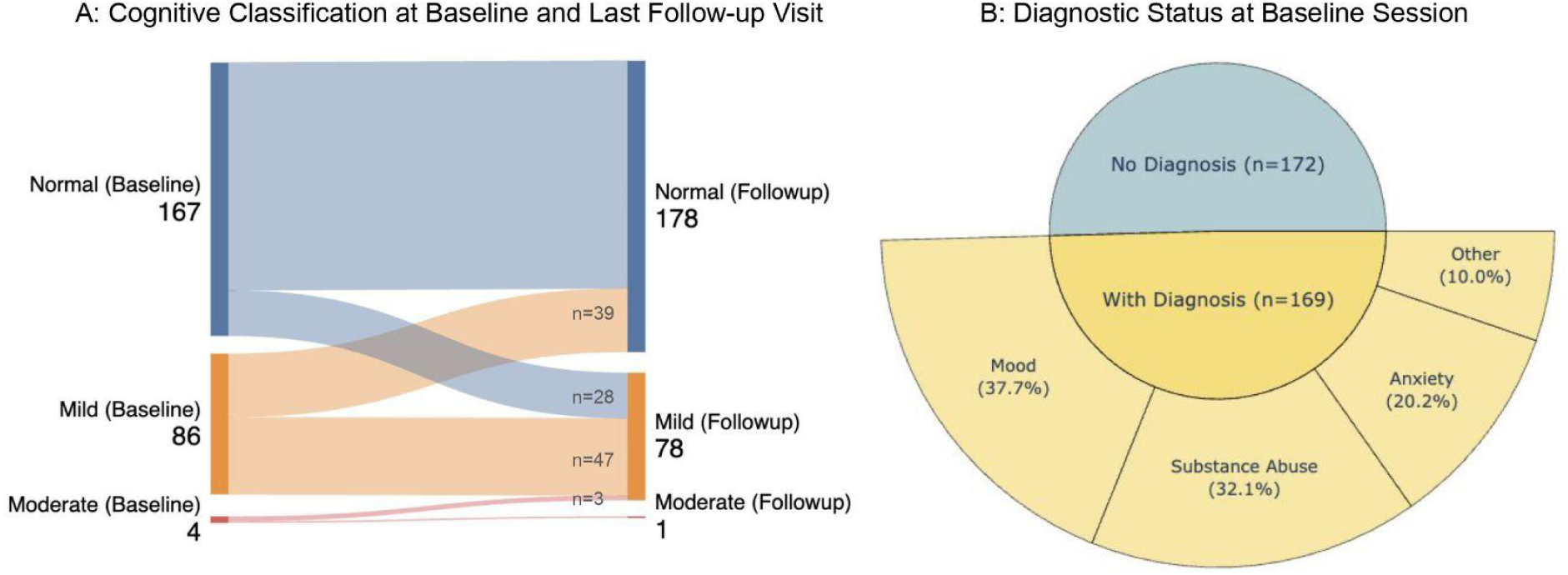
Diagnostic classifications of cognition and major psychiatric categories. A. Change in cognitive classification based on MoCA for all participants with baseline and at least one follow-up visit (n = 257). Scores at baseline (left) are compared with the last follow-up session for each participant (right). No follow-up information was available for 86 participants classified “normal” (n= 56) and “mild impairment” (n= 30) at baseline. B. DSM-IV diagnostic status at baseline (via Structured Clinical Interview for DSM-IV-TR Axis I Disorders – Non-Patient Edition (SCID-I/NP),^29^ n = 341) and the relative percent of mood, substance, anxiety, and other diagnoses. Includes past and present DSM-IV diagnoses. “Other” diagnoses included: Hypochondriasis: n = 2; Anorexia Nervosa: n=5; Eating Disorder NOS: n=6; Bulimia Nervosa: n=3; Attention-Deficit/Hyperactivity Disorder: n=2; ADHD, Predominantly Inattentive Type: n=2; ADHD, Combined Type: n=1; ADHD NOS: n=1; Bereavement: n=4. Multiple diagnoses per participant were low (two = 42, three = 19, four or more = 17) and not included in the figure.

Psychiatric characterization via semi-structured SCID-I/NP interviews determined approximately half of the participants (n = 172, 51%) had no DSM-IV diagnosis at baseline. Among those with a diagnosis (n = 169, 49%), major diagnostic categories were relatively evenly split among mood (38%), substance abuse (32%), and anxiety (20%). There was a relatively low frequency of other disorders (10%) and multiple diagnoses per participant (21% with three or more diagnoses). See Figure 3b for more detail.

### Quality assessment

#### Phenotypic data

Prior to each NKI-RS release, scatterplots by age and boxplots comparing past releases to each progressive release were reviewed. Phenotypic data were not “cleaned” of outliers unless overt technical errors were identified in pre-release reviews. Data review and cleaning is expected to be incorporated into each independent research analysis.

Figures 4–6 summarize score distributions and age-associated performance trends for core cognitive and mental health measures. Figure 4 illustrates both roughly symmetric (e.g., WASI-II index scores, WIAT-II Numerical Operations) and skewed distributions (e.g., WIAT-II Reading and Spelling, MoCA, BDI, STAI). Figure 5 shows modest but statistically significant age effects for the Attention Network Task in alerting, conflict, and grand mean reaction time. Figure 6 (top row) demonstrates age-related performance declines on D-KEFS Category Fluency, Trail Making Test–Switching, and Color Word Interference–Inhibition. Figure 6 (bottom row) highlights relatively stable performance across age for Letter Fluency, Trail Making Test–Sequencing, and a composite score derived from the baseline conditions of the Color Word Interference Test (Color Naming and Word Reading). Greater variability in performance was observed for conditions with longer completion times (Trail Making Test–Switching, Color Word Interference–Inhibition) and a greater number of total words generated (Category Fluency).

**Figure 4.**
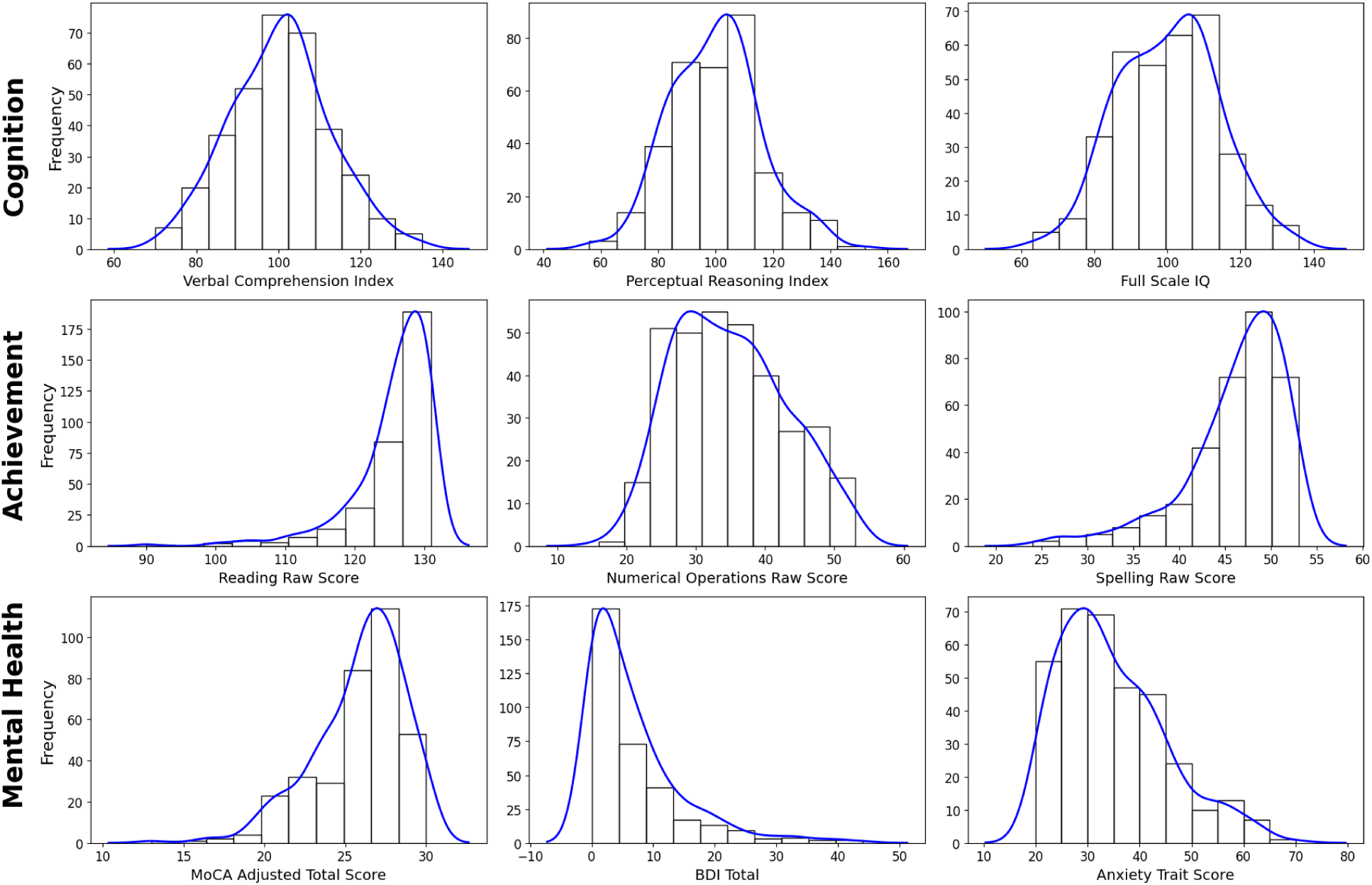
Distributions of cognitive abilities, academic achievement, and mental health measures. *Top row*: composite scores from WASI-II. *Middle row*: raw scores from WIAT. *Bottom row left to right*: MoCA education-adjusted total score, BDI total score, and STAI raw Anxiety trait score. Only baseline data are included and no filtering was done.

**Figure 5.**
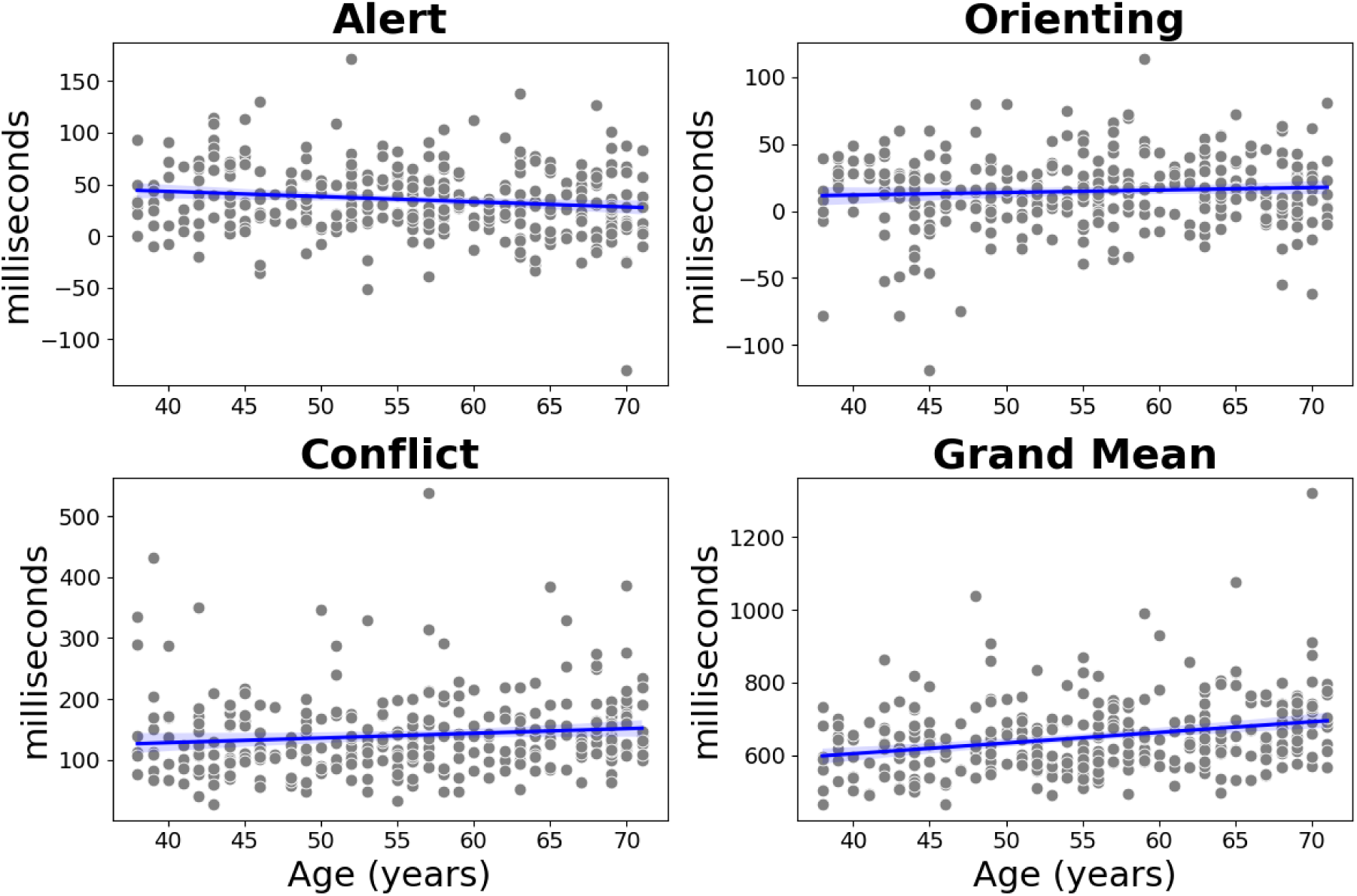
Scatter plots of Attention Network Task subscores and grand mean against age. Alert: t(332) = -2.73, p = .007; Orienting: t(332) = 1.23, p = .220; Conflict: t(330) = 2.12, p = .034; Grand Mean: t(330) = 5.33, p<.001. Plots include all unfiltered baseline data.

**Figure 6.**
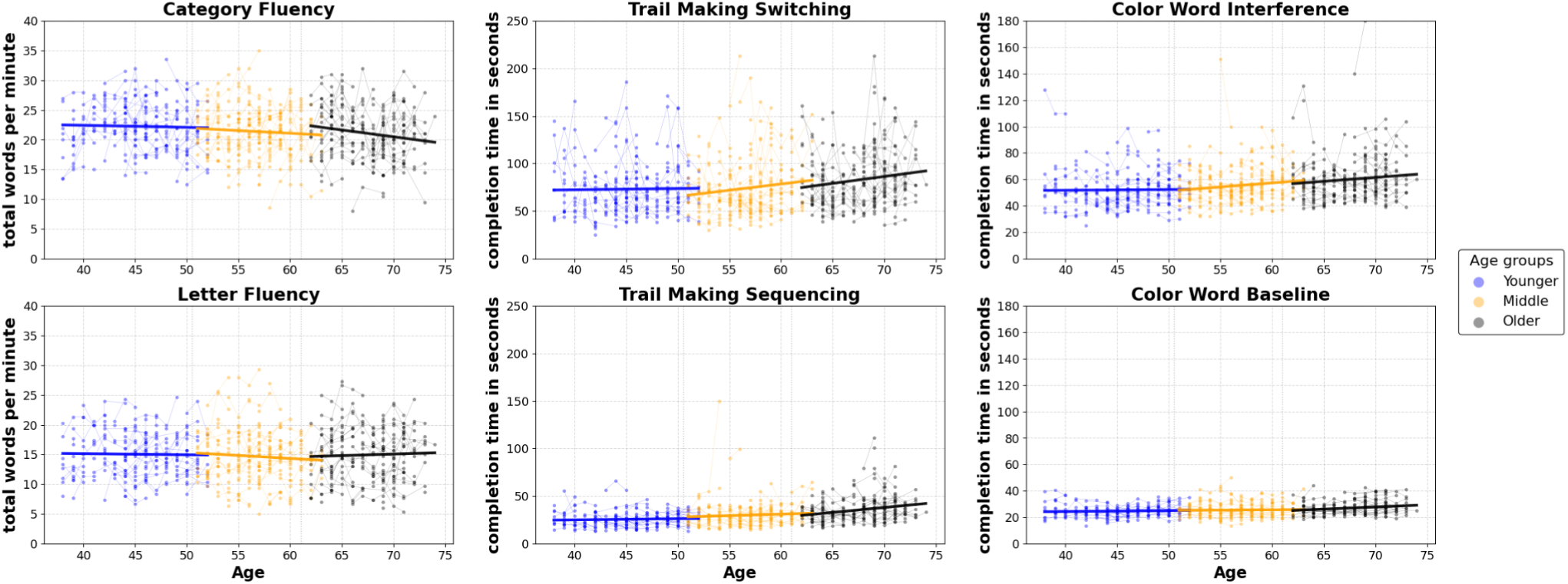
Delis-Kaplan Executive Function Scale Subtest Scores across longitudinal timepoints, color coded for three age tertile groups (Younger (blue): <=50.5; Middle (orange): >50.5 and <= 61; Older (black): > 61). Left panel shows performance on category and letter fluency (CF, LF). Middle panel shows Trail Making Test switching and sequencing conditions (Switch, SeQ). Right panel shows Color Word Interference Test interference and averaged baseline conditions (INT, BASE). Three bolded lines are regression lines for the three age groups in each plot. Significant age effects were observed for: CF Older: t(256) = −2.60, p = .010; Switch Middle: t(263) = 2.10, p = .037; Switch Older: t(263) = 2.43, p = .016; SeQ Older: t(280) = 4.15, p < .001; INT Middle: t(268) = 2.23, p = .026; BASE Older: t(270) = 3.33, p = .001.

Figure 7 presents the histogram and kernel density estimate for distributions of physical health measures, including grooved pegboard, grip strength, walk time, and VO₂ max.

**Figure 7.**
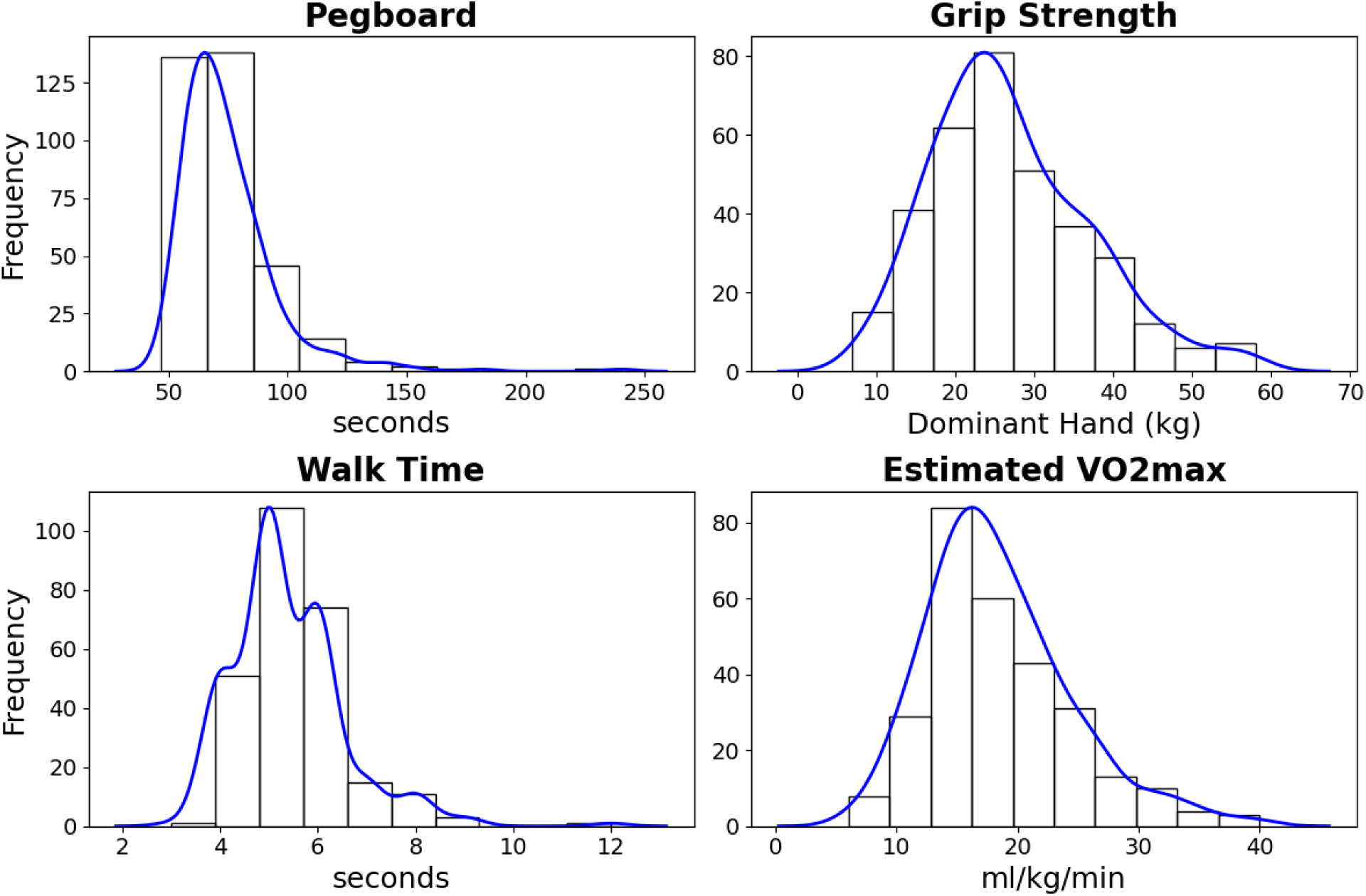
Distributions of physical measures: *Top: left* - Grooved pegboard completion time for dominant hand, *right* - Grip strength for dominant hand. *Bottom*: *left* - Single task walking time from Walking While Talking Task, *right* - Estimated VO_2_max from submaximal cardiorespiratory fitness test. Only baseline data are included and no filtering was done.

Supplementary Figure 2 depicts boxplots with kernel-density overlays for each analyte from the blood sample collection. Distributions were largely unimodal and centered within expected adult ranges, with several measures showing right-tailed skew typical of metabolic/inflammatory indices. Extreme values are visible in the tails; consistent with our phenotypic-data policy, these observations were retained (no outlier trimming) to maximize transparency and enable users to apply domain-appropriate handling (e.g., robust estimators, winsorization) in downstream analyses. Missingness was modest and chiefly attributable to occasional deferred phlebotomy or scheduling constraints. See also Supplementary Table 3 for a summary by assay (sample sizes, median, IQR, 5th–95th percentiles).

To evaluate the long term stability of key phenotypic measures over time, intraclass correlation coefficients (ICCs) were calculated using participants with at least three annual longitudinal assessments. Cognitive indices from the WASI-II showed good stability, with agreement/consistency values ranging from .67/.71 (Similarities) to .77/.77 (Matrix Reasoning) to .79/.83 (Vocabulary) to .90/.90 (Block Design). The MoCA demonstrated moderate stability (.63/.64), while affective measures were higher (Beck Depression Inventory; BDI = .75/.75; State Trait Anxiety Inventory-Trait; STAI-T= .81/.81). Practice-related gains were observed for the WASI-II and MoCA, whereas BDI and STAI-T scores remained stable (See Supplementary Table 4 for details here and below). Estimates for the Attention Network Task, consistent with prior reports,^30^ showed moderate test-retest stability for conflict (.48/.50) and grand mean reaction time (.66/.67), but lower values for alerting (.38/.39) and orienting (.16/.17). Practice effects included increases in alerting and orienting effects, decreases in conflict effects, and faster overall response times across sessions. D-KEFS tasks demonstrated moderate to high retest stability, with Color Word Interference scores ranging from .85 to .87 across baseline and interference conditions. Trail Making Test sequencing conditions showed moderate retest stability (number = .57/.58; letter = .64/.65), while the switching condition was higher (.74/.75) and most sensitive to practice effects, t(196) = −2.78, p=.006. Tower total achievement score showed moderate retest stablility (.53/.55) with improved scores over time (t(197)= 6.88, p <.0001, and verbal fluency measures were higher (letter = .83/.84; category = .72/.73) with increasing word production across sessions, t(177) = 4.27, p<.001 and t(178)=2.60, p=.01 for letter and category, respectively. Motor praxis reaction time, conditional exclusion test reaction time, two-back total correct, and continuous performance test d-prime from the Penn Computerized Neurocognitive Battery (Penn CNB)^31^ ranged in test-retest stability from a low of .47 consistency for two-back to a high of .74 agreement for the continuous performance test. Performance gains over annual follow-up visits were observed for motor praxis, two-back, and the continuous performance test, but not for reaction time on the conditional exclusion test (see Supplementary Table 4). Physical measures ranged from relatively high test-retest stability (grip strength = .78/.78; VO₂ max = .83/.83) to moderate (pegboard completion = .68/.69; walk time = .46/.47). There were no observed practice effects for grip strength or VO₂ max, while pegboard times decreased with repeated testing and walk times increased. Overall, most measures demonstrated moderate to high longitudinal stability, with practice effects most pronounced in cognitive and motor speed tasks. ICCs and mean slopes for a sampling of the measures are reported in detail in Supplementary Table 4.

We next examined correlations among phenotypic variables to facilitate evaluation of measures (Figure 8). All correlations were corrected for false discovery rate. To ensure transparency, outliers were not removed for this descriptor, though we recommend careful data review and outlier handling in subsequent analyses. Broad patterns of association across cognitive, health, and demographic domains were consistent with established findings^32,33^. For example, (1) age correlated with fluid (BD, MR) but not crystallized (VOC,SIM) WASI-II subtests^34–36^,was negatively associated with RAVLT delayed recall^37,38^ and category fluency,^39,40^ and positively associated with Trail Making Test completion time;^40–42^ (2) age was related to poorer physical but not mental health^43–45^; (3) self-reported mood, sleep, and subjective cognitive impairment were intercorrelated;^46,47^ (4) the Attention Network Task conflict effect and grand mean response times correlated with other cognitive measures, whereas alerting and orienting did not;^48^ (5) the MoCA was associated with episodic memory,^49^ executive function,^50^performance on WASI-II subtests,^51^ academic achievement,^52^ and physical health measures (BMI,^53^ pegboard,^54^ walk time,^23,54–56^ VO₂max^6^), but not with age^21,57^ or subjective complaints;^58^ and (6) VO₂max cardiorespiratory fitness was correlated with all measures of cognition except ANT alerting and orienting^59,60^, social network size^61^, and grip strength^62^, and was negatively associated with BMI^62^ and blood glucose^63^. With respect to MoCA performance and age, some prior reports did not find an association for age in healthy adults between 55 and 85 years old^21^, while others did.^64^ Age cohort differences in education level may also be an important factor to consider.^65^ See Supplementary Figure 4 for MoCA scatterplots by age. See also, Supplementary Figure 3 for additional correlations between participants’ demographic and phenotypic characteristics split by age 65 and older (OASR) and under 65 (ASR).

**Figure 8.**
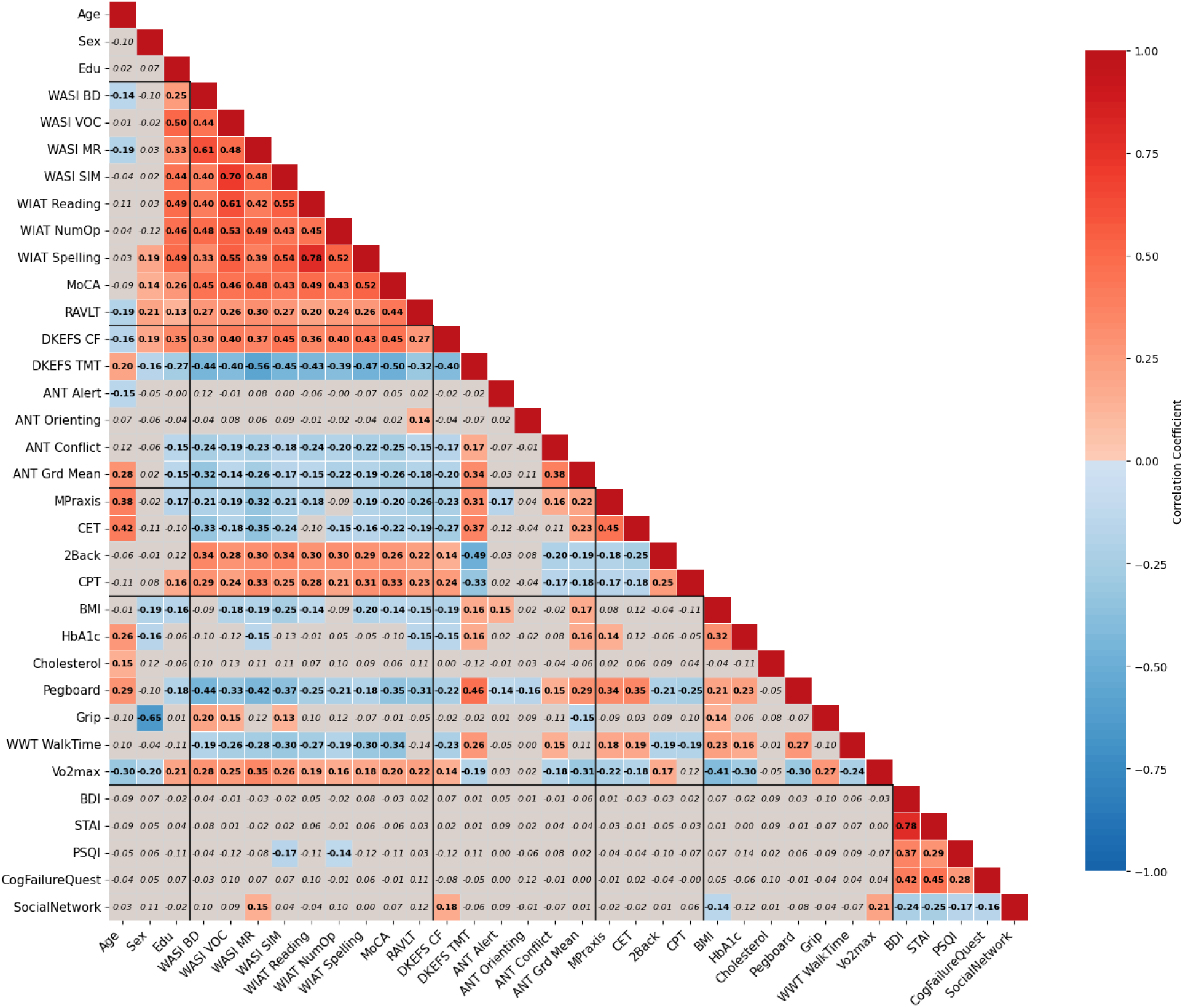
Heat map showing r values for bivariate correlations among demographic and phenotypic measures. Values that did not survive False-Discovery Rate multiple-comparisons correction are shown in gray. Four correlations with the most missing data were: WWT WalkTime & HbA1c (missing n=136), WWT WalkTime & Cholesterol (missing n= 132), HbA1c & VO_2_max (missing n= 118), and Cholesterol & VO_2_max (missing n=113). *WASI BD*: Wechsler Abbreviated Scale of Intelligence Board Design; *WASI VOC*: Vocabulary; *WASI MR*: Matrix Reasoning; *WASI SIM:* WASI Similarities; *WIAT Reading:* Wechsler Individual Achievement Test Word Reading Raw Score; *WIAT NumOP*: Numerical Operations Raw Score; *WIAT Spelling*: Spelling Raw Score; *MoCA*: Montreal Cognitive Assessment total score adjusted for education; *RAVLT:* Rey Auditory Verbal Learning Test - Long-Delay Recall; *DKEFS CF*: Delis-Kaplan Executive Function System Category Fluency Raw Score; *DKEFS TMT*: DKEFS Trail Making Test; *ANT Grd Mean*: Attentional Network Test - Grand Mean; *MPraxis*: PENN Computerized Neurobehavioral Battery (CNB) - Motor Praxis Reaction Time; *CET*: PENN CNB Conditional Exclusion Task RT; *2Back*: PENN CNB 2-Back Total Correct; *CPT*: PENN CNB Continuous Performance Test Number + Letter d-prime; *BMI*: Body Mass Index; *HbA1c*: Hemoglobin A1c; *Grip*: Grip Strength in kg for dominant hand; *WWT WalkTime*: Walking While Talking - First trial of single task walking time; *VO_2_max*: estimated from the submaximal test; *BDI*: Beck Depression Inventory Total; *STAI*: State-Trait Anxiety Inventory - Total trait raw score; *PSQI*: Pittsburgh Sleep Quality Index total sleep score; *CogFailureQuest*: Cognitive Failures Questionnaire Total Score; *Social Network*: Social Network Size. Only baseline data are included and no filtering was done.

### Neuroimaging data

All NKI-RS imaging datasets are released to users without quality filtering, as there is no universal agreement on quality standards in the field. Additionally, including datasets of varying quality serves two important purposes: it enables researchers to develop new methods for artifact correction and allows for assessment of how real-world data imperfections affect reliability and reproducibility of findings.

### Structural MRI

T1-weighted images were processed with Mindboggle^66^, a software that combines functionalities from Advanced Normalization Tools (ANTS)^67^ and FreeSurfer^68^ to parcellate brain regions using the Desikan-Killiany-Tourville atlas.^69^ Euler number was calculated in FreeSurfer to assess data quality, with poorer quality indicated by more negative values.^70^ Out of 975 T1 images across all timepoints, 4.0% had Euler numbers more than 2SD below the mean. As shown in Figure 9, Euler numbers correlated negatively with age (r = −.196, p<.001) and, except for dMRI, also correlated with FD from all other sequences (r between −.165 to −.363, all p < .01).

**Figure 9.**
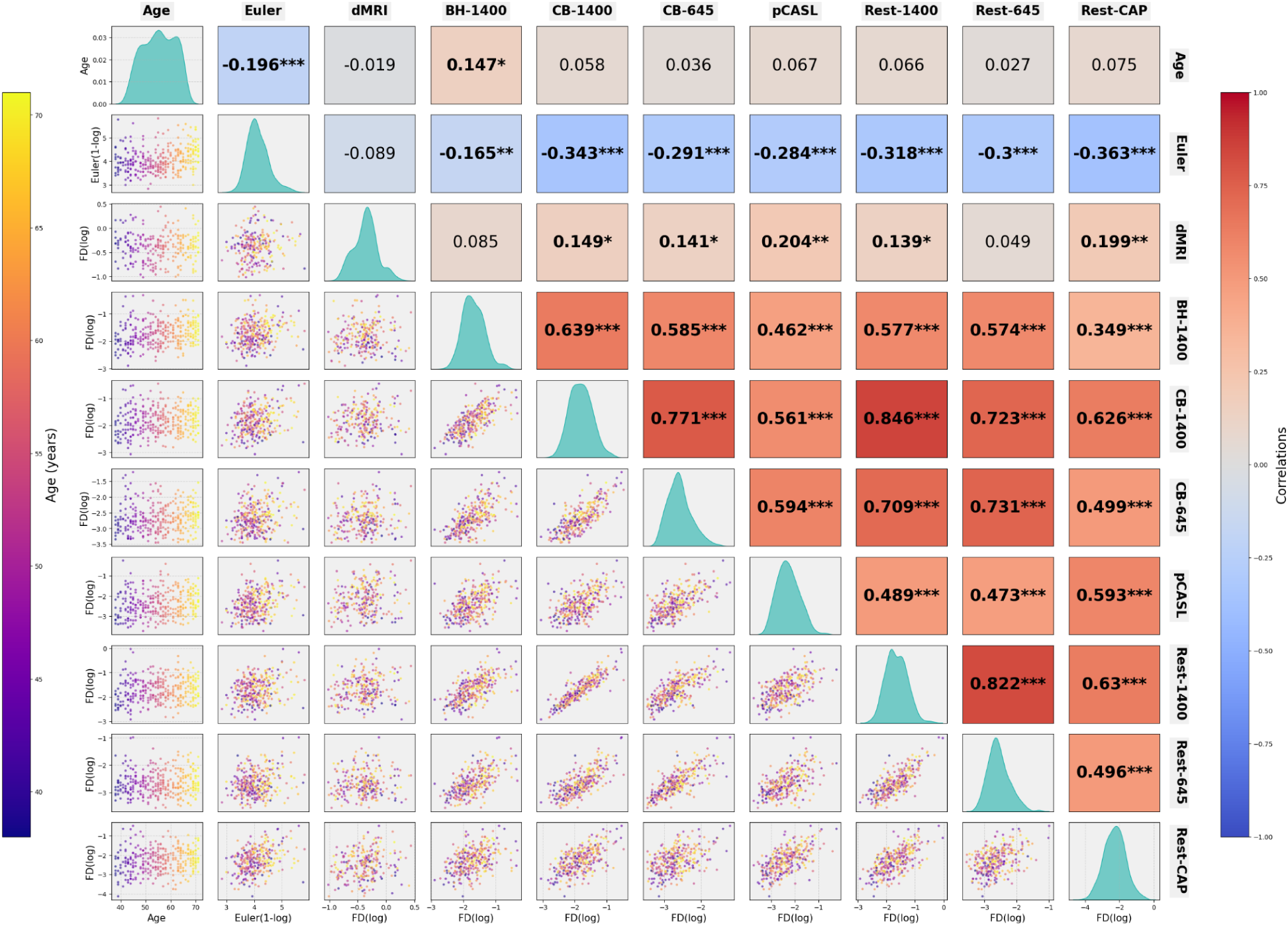
Correlations of mean Framewise Displacement (FD) for MRI sequences with age and FD for other sequences. Lower triangle shows scatterplots with log-transformed FDs, and points were color-coded by participant age. Upper triangle displays the corresponding Pearson correlation coefficients (r) for each correlation, with significance indicated by asterisks (*p < 0.05; **p < 0.01; ***p < 0.001) based on FDR-corrected p-values. Diagonal panels show the distribution of FD values for age and each sequence.

### Diffusion MRI

Diffusion MRI data were processed in QSIPrep and QSIRecon.^71^ Detailed description of the full pipeline is described in Supplementary Information. Briefly, QSIPrep performed standard preprocessing using MRtrix3 tools,^72^ including denoising, Gibbs unringing, and bias correction. Head motion and Eddy current corrections were then performed with FSL’s eddy tool.^73^ Average Framewise Displacement (FD) and age effects are shown in Figure 9. After preprocessing, QSIRecon’s autotrack pipeline^74^ was used to reconstruct 56 white matter tracts and mean microstructural measures (FA) were calculated for each tract and averaged between the two hemispheres. With a total of 839 dMRI images, 3.34% had FD larger than 2 SD from the mean and 68% of the scans had no bad slices. There was no correlation between age and FD. Age differences in FA for the whole brain and select white matter tracts are presented in row 3 of Figure 10. Consistent with current literature, FA declines in older age and at the steepest rate in the oldest age group.

**Figure 10.**
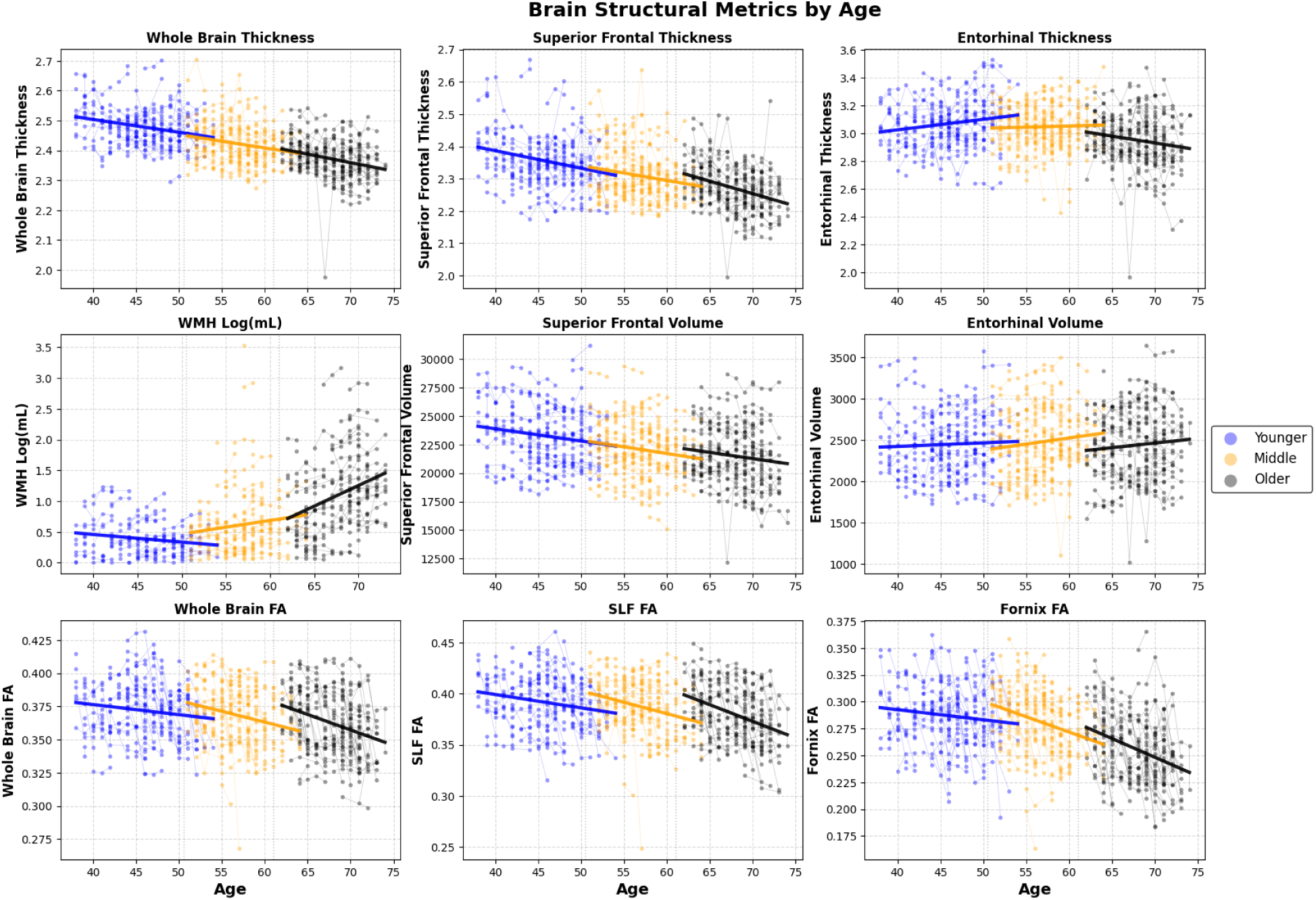
Structural measures from MRI for individual participants collapsing across all timepoints and color coded for three age tertile groups (Younger: <=50.5; Middle: >50.5 and <= 61; Older: > 61). Row 1 shows the mean cortical thickness for the whole brain and for the Superior Frontal and Entorhinal Cortices (WBt, SFt, ERt). Row 2 shows White Matter Hyperintensity and volumes for the Superior Frontal and Entorhinal Cortices (WMH, SFv, ERv). Row 3 shows the mean fractional anisotropy (FA) across 28 major white matter tracts, for the Superior Longitudinal Fasciculus, and for the Fornix (WBfa, SLFfa, Fxfa). Three bolded lines are age regressions in each plot. Significant age effects were observed for: WBt - Younger: t(282) = −4.25, Middle: t(291) = −3.46, Older: t(313) = −5.09, all p<.0001; SFt - Younger: t(282) = −4.34, p < .0001, Middle: t(291) = −3.15, p = .0018, Older: t(313) = −6.14, p<.0001; ERt - Younger: t(282) = 2.81, p = .0053, Older: t(313) = −2.91, p = .0039; WMH: Younger: t(196) = −2.28, p = .0238, Older: t(252) = 5.28, p<.0001; SFv: Younger: t(282) = −2.70, p = .0074, Middle: t(291) = −2.50, p = .0131, Older: t(313) = −2.25, p = .0251; WBfa: Younger: t(263) = −2.16, p = .0314, Middle: t(264) = −3.63, p = .0003, Older: t(290) = −5.21, p <.0001; SLFfa: Younger: t(263) = −3.06, p = .0024, Middle: t(264) = −4.25, Older: t(290) = −6.37, both p<.0001; Fxfa: Middle: t(263) = −4.92, Older: t(290) = −6.02, both p<.0001.

### Functional MRI

Jenkinson’s FD^75^ was calculated for task and resting fMRI scans using AFNI’s 3dvolreg function.^76^ Figure 9 shows the cross-correlations among mean FDs for fMRI scans and each with age and other sequences. Among the six fMRI sequences, out of a range of 684 to 696 total images across all timepoints, the percent of data with FD greater than 2 SD than the mean was between 2% to 4%. Among all fMRI sequences, only FD for the Breath-hold sequence with TR of 1400ms was correlated with age.

Along with other NKI-RS datasets, preprocessed fMRI, T1 and T2-weighted structural MRI images from MIVAC are provided through the Reproducible Brain Charts (RBC) project (https://reprobrainchart.github.io/). Data in RBC was preprocessed using the Configurable Pipeline for the Analysis of Connectomes (C-PAC).^77^ Details about preprocessing are provided in.^78^

### Pseudo-Continuous Arterial Spin Labeling (pCASL)

FD for pCASL data were quantified with AFNI’s 3dvolreg function. WIth a total of 680 images across all timepoints, 3.4% of participant data exceeded 2 SD above the mean FD across subjects.

### Fluid-attenuated inversion recovery (FLAIR) MRI

FLAIR data were segmented into normal and hyperintense regions with the Lesion Growth Algorithm from the Lesion Segmentation Toolbox (LST v3.0; ^79^) with a kappa threshold of 0.3. As expected, Figure 10 shows that the oldest age group follows the steepest increase in total WMH volume.

## USAGE NOTES

The MIVAC dataset extends the NKI-RS initiative into midlife and older adulthood, offering a deeply phenotyped, openly available resource for examining normative and atypical trajectories of brain aging. The dataset is designed for broad application across neuroscience, psychology, gerontology, and public health, among other fields. Since its inception, the NKI-RS has supported the training of emerging scientists at the undergraduate, master’s, and doctoral levels, as well as advanced research by established investigators—accelerating discovery and fostering the next generation of basic and clinical research. Below we outline key considerations for effective use.

### Data access and formats

All imaging data are distributed in BIDS format and can be downloaded directly via Amazon S3, using either bulk download scripts or open-source clients. Phenotypic and physiological data are available via the LORIS and COINS platforms, with different access levels depending on whether users request the full phenotypic release (requiring a DUA) or the Lite release (limited phenotyping, no agreement required).

### Data quality

In keeping with NKI-RS open science principles, all data are released without exclusion on the basis of quality. This allows researchers to apply their own quality-control thresholds and to develop or benchmark artifact-correction methods. Imaging data include quality metrics such as framewise displacement and Euler numbers to aid downstream decisions. Phenotypic data are released without removal of outliers, with the expectation that users apply study-specific cleaning and screening procedures appropriate to their research aims.

### Longitudinal considerations

The MIVAC substudy employed a multi-cohort longitudinal design, with repeated measures up to four years. Due to the COVID-19 pandemic, data collection was interrupted during the third annual follow-up period, resulting in substantial missingness for the 4th timepoint. Variables coding visit intervals (e.g., “day lag”) are provided to allow precise modeling of within-person change and practice effects. Users should account for attrition and missing data when conducting longitudinal analyses.

### Phenotypic protocols

Measures were harmonized across prior NKI-RS substudies to facilitate lifespan analyses. Some measures (e.g., CASI-A, MoCA) were extended to broader age ranges and include item level data for methodological analyses. Users are advised to consult the full End User Protocol (https://rocklandsample.org/mivac-full-end-user-protocol-2) and detailed item level information in a downloadable file here: https://rocklandsample.org/for-researchers/rockland-sample-i-2011-2022/assessments/assessments_documentation.

### Cardiorespiratory fitness and physical health measures

This dataset incorporates gold-standard submaximal VO₂max assessments, providing an opportunity to examine modifiable protective factors in aging. Investigators should review cardiorespiratory assessment quality flags when incorporating this assessment into analyses. For blood-based analyte analyses, we recommend (i) log-transforming long-tailed biomarkers, (ii) explicitly modeling fasting status and relevant medications where available, and (iii) reporting both robust (median/IQR) and parametric (mean/SD) summaries.

### End user support

An established inquiry mechanism is available to assist with questions about access, documentation, and data interpretation. Inquiries are reviewed by an End User Response Panel composed of investigators with expertise in imaging, phenotyping, and data management.

### Intended use

The dataset is well-suited for studies of:

- normative and atypical trajectories of brain aging beginning in midlife,
- the role of modifiable health factors (e.g., cardiorespiratory fitness, BMI, sleep, social networks) in cognitive and neural outcomes,
- methods development for artifact detection, denoising, or harmonization across open imaging datasets, and
- lifespan analyses leveraging harmonized protocols across NKI-RS substudies.

### Limitations

Limitations include incomplete longitudinal follow-up due to pandemic-related shutdown, uneven representation of sex (female > male), and the need for users to implement quality-control and outlier handling. Researchers should interpret findings within the context of these constraints.

## DATA AVAILABILITY

Information for researchers about the Rockland Sample study and how to obtain data can be obtained in the study website: https://rocklandsample.org/for-researchers

Phenotypic data is available in two platforms:

- LORIS: https://data.rocklandsample.rfmh.org
- COINS: https://coins.trendscenter.org

We recommend users obtain data through the LORIS portal, since it contains updated study codes (https://rocklandsample.org/for-researchers-rockland-sample-i-2011-2022-documentation-study-codes).

Step-by-step instructions on how to download neuroimaging data can be found here: https://rocklandsample.org/accessing-the-neuroimaging-data-releases

Neuroimaging data is hosted on an open AWS S3 bucket. BIDS organized data can be directly accessed through this S3 path: s3://fcp-indi/data/Projects/RocklandSample/RawDataBIDSLatest

### Data Citation

*Enhanced Nathan Kline Institute - Rockland Sample. International Neuroimaging Data-Sharing Initiative* https://doi.org/10.15387/fcp_indi.retro.NKIRockland *(2013)*.

## CODE AVAILABILITY

No custom codes or algorithms were used to generate or process the data presented in this manuscript.

## ACKNOWLEDGMENTS

We thank the research participants for their generous contributions of time and effort. We appreciate the study team members who supported data collection, data management, preliminary analyses, literature reviews, community outreach, and recruitment: Julia Beatini, Brian Bengyak, Sinead Burrows, Jerlyne Calixte, Brian Carbone, Stephanie Carelli, Galen Cassidy, Maya Charan, Jessica Cloud, Jesenya DeLeon, Lauren Futterman, Gwen Geisler, Chelsea Gessner, Alyssa Giannone, Jamie Glass, Courtney Gray, Steven Homan, Olive Hwang, Christy Joseph, Stephanie Kamiel, Alexis Lieval, George Lopez, Bryanna Mackey, Amalia McDonald, Laura Panek, John Pellman, Hayley Reed, Margaret Ryan, Sheela Sajan. We thank the CBIN Design, Acquisition, and Neuromodulation Laboratory’s MR technical staff Caixia Hu and Raj Sangoi, Tina Bermudez, Kefin Sajan of the CBIN Computational Neuroimaging Laboratory for his neuroinformatics support, and Elizabeth Zakszewski and Gabriel Schubiner of the Child Mind Institute (CMI). We also thank Nathalia Esper for image preprocessing support. We thank Amazon Web Service (AWS) for hosting our neuroimaging data through their Open Data Sponsorship Program. The Mapping Interindividual Variation in the Aging Connectome was principally supported by NIH R01AG047596 (MPIs Colcombe/Milham). A subset of participants had baseline characterizations supported by the core enhanced NKI-RS protocol NIMH BRAINS R01MH094639-01 (PI Milham) and NIH R01MH101555 (PI Craddock). Funding for key personnel was also provided in part by the New York State Office of Mental Health and Research Foundation for Mental Hygiene.

## AUTHOR CONTRIBUTIONS

Conception and Experimental Design: S.J.C., M.P.M., A.M-B., V.G., R.H.T. Implementation and Logistics: S.J.C., M.P.M., A.M-B, R.H.T., M.B., M.K., K.D.T. Data Collection: M.B., M.K., A.M-B., R.H.T., K.D.T. Data Informatics: A.R.F., D.G-B, Y.G., L.G., S.J.C., O.R., M.K., M.B., A.M-B, K.X.G., Data Analysis: S.J.C., A.M-B., D.G-B, Y.G.,L.G., O.R.,V.G., A.R.F. Initial Drafting of the Manuscript: A.M-B., S.J.C., A.R.F., K.D.T., O.R., K.X.G. Critical Review and Editing of the Manuscript: All authors contributed to the critical review and editing of the manuscript.

## SUPPLEMENTARY MATERIALS

### Supplementary Figures

**Figure S1.**
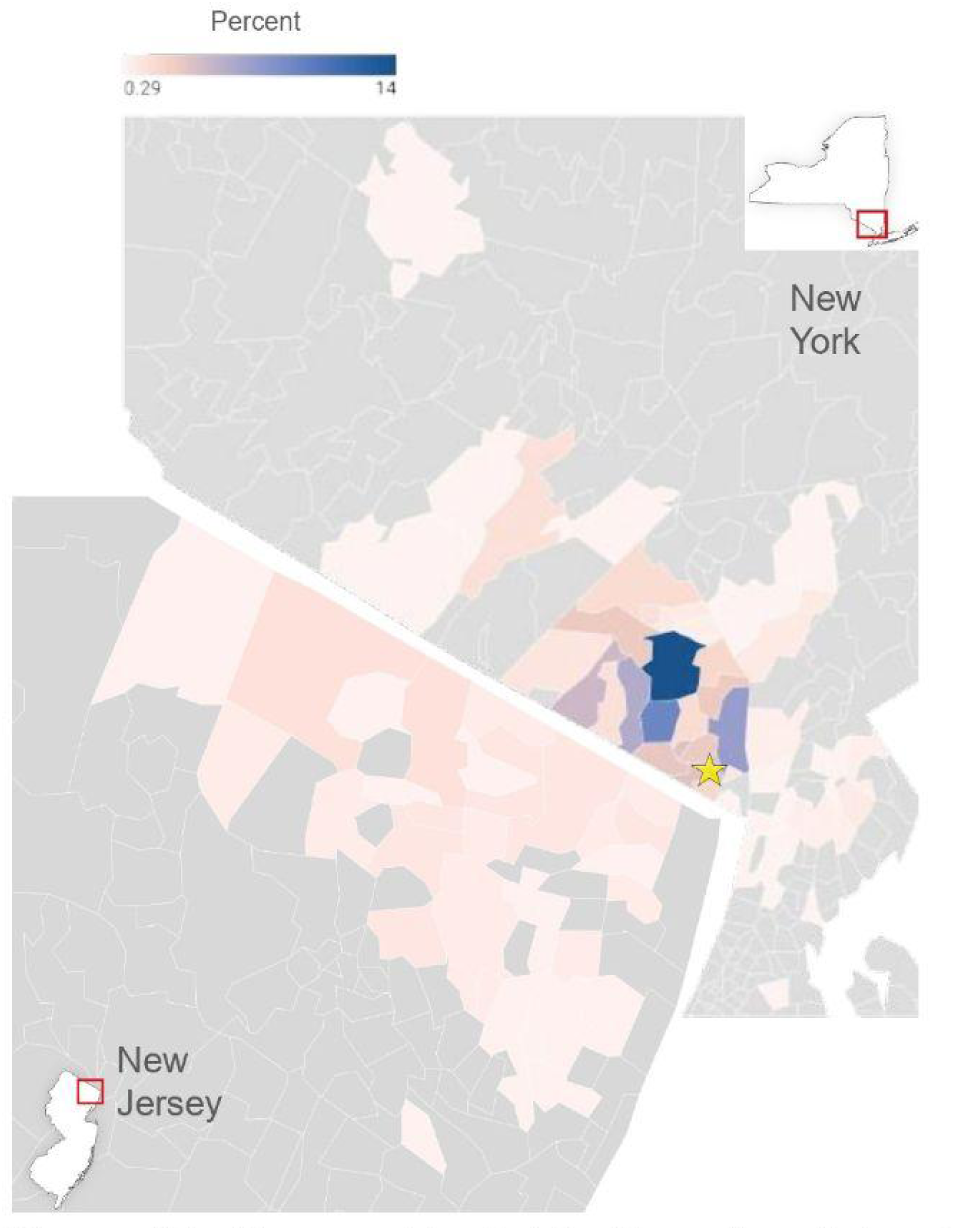
Geographic distribution of participants by zip code. Color indicates percentages of participants in each zip code. Star indicates location of the Nathan Kline Institute.

**Figure S2.**
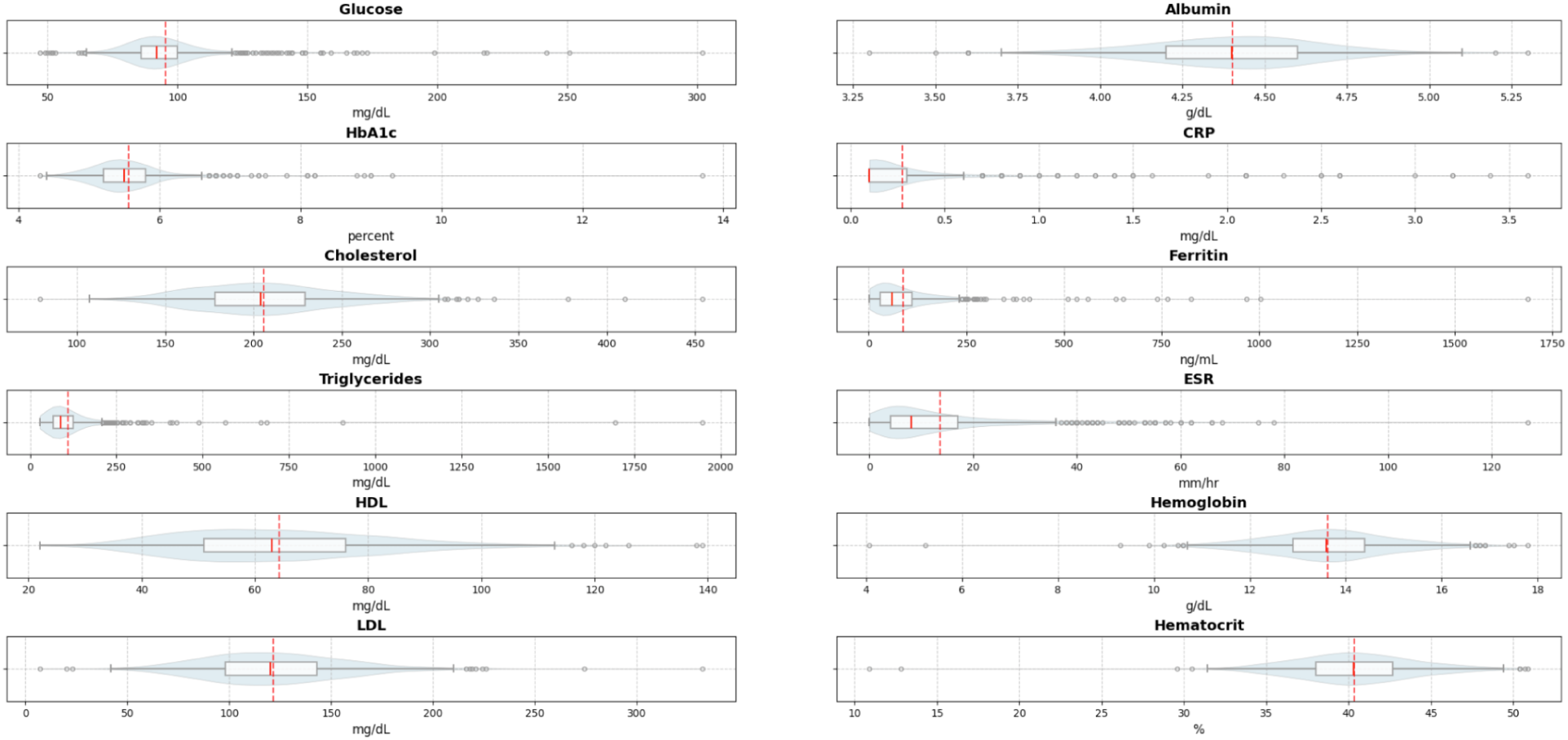
Boxplots with kernel-density overlays for each analyte from the blood sample collection. HbA1c = Hemoglobin A1C; HDL = High-density lipoprotein; LDL= Low-density lipoprotein; CRP = C-reactive protein; ESR = Erythrocyte sedimentation rate.

**Figure S3.**
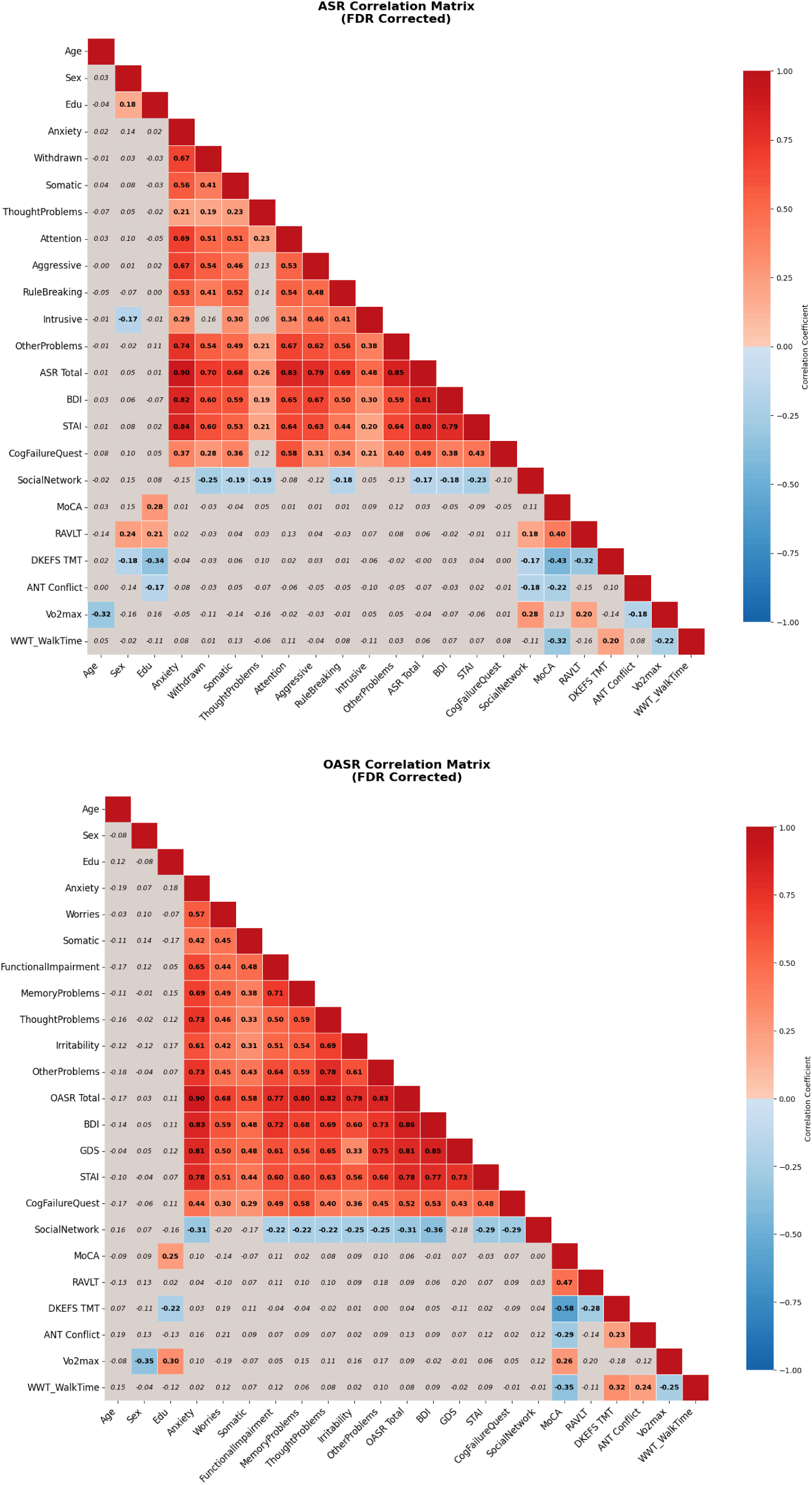
Heat map showing coefficients for bivariate correlations among demographic and subset of phenotypic measures against subscales of Adult Self Report (ASR; administered to participants <60 years old) and Older Adult Self Report (OASR; administered to participants 60+ years old) and total score. Values that survived False-Discovery Rate multiple-comparisons correction are shown in bolded font. *BDI*: Beck Depression Inventory Total; *GDS*: Geriatric Depression Scale; *STAI*: State-Trait Anxiety Inventory - Total trait raw score; *CogFailureQuest*: Cognitive Failures Questionnaire Total Score; *Social Network*: Social Network Size, *MoCA*: Montreal Cognitive Assessment total score adjusted for education; *RAVLT*: Rey Auditory Verbal Learning Test - Long delay recall; *DKEFS TMT:* Delis-Kaplan Executive Function System Trail Making Test; *ANT*: Attention Network Test; *VO2max*: estimated from the submaximal test; *WWT WalkTime*: Walking While Talking - First trial of single task walking time. Only baseline data are included and no filtering was done.

**Figure S4.**
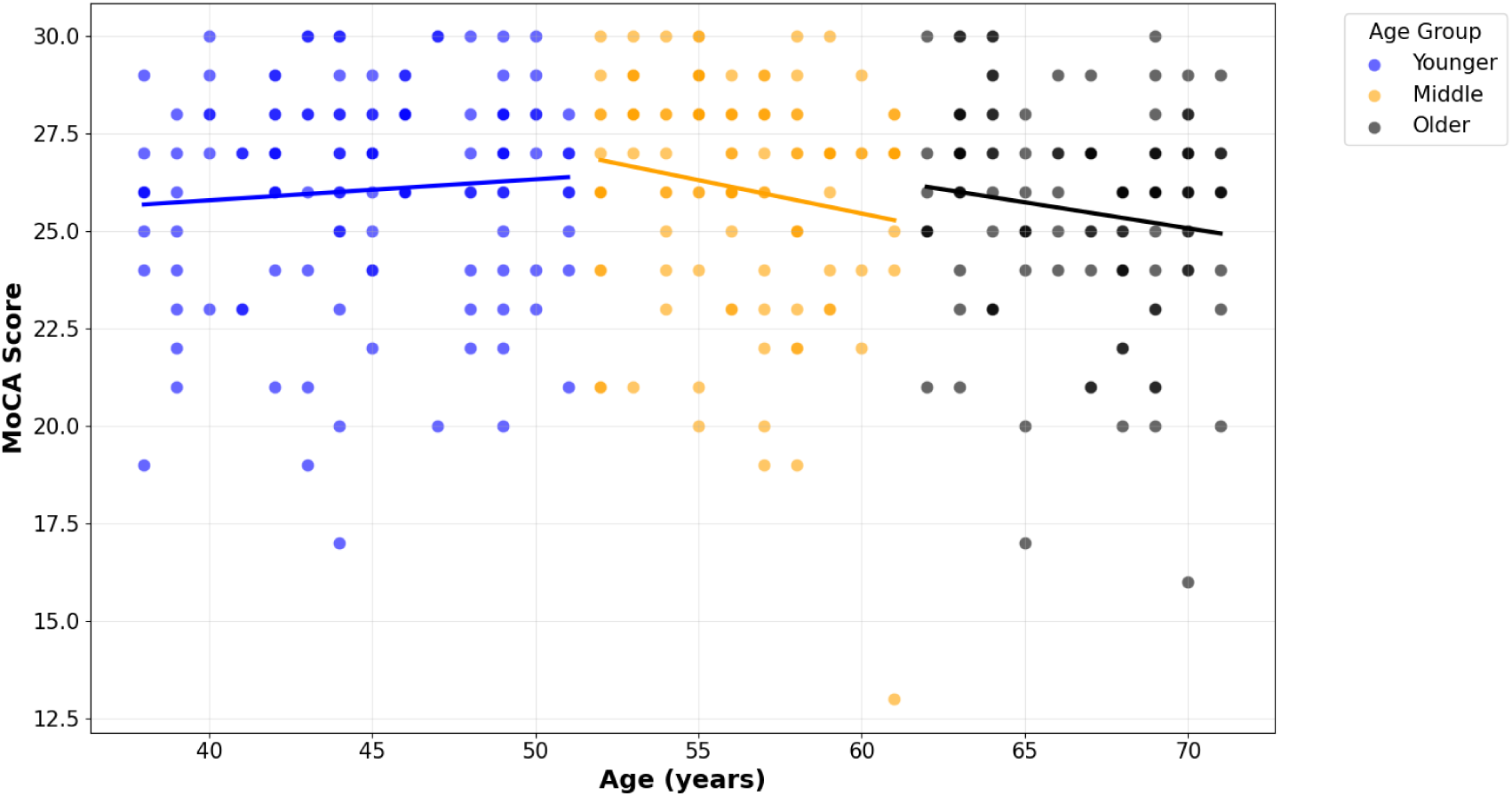
Scatter plot of MoCA education adjusted total score against Age, divided into three age groups by tertiles (Younger: </= 51; Middle: > 51 and </= 61; Older: > 61). The three regression lines are specific to each age group (Younger: t (119)= 0.83, p = 0.41; Middle: t(108) = −1.66, p = 0.10; Older: t(110) = −1.49, p = 0.14). All unfiltered baseline data were included in the plot.

### Supplementary Tables

**Table S1a.** Mapping of Older Adult Self Report (OASR) and Adult Self Report (ASR) common items and subscales.

| Question | ASR # | OASR # | Question | ASR # | OASR # |
| --- | --- | --- | --- | --- | --- |
| **Somatic Complaints** |  |  | **Anxious/Depressed** |  |  |
| Dizzy | 51 | 46 | Lonely | 12 | 11 |
| Aches | 56a | 49a | Cries | 14 | 13 |
| Headaches | 56b | 49b | Worries future | 22 | 21 |
| Nausea | 56c | 49c | Fears doing bad | 31 | 28 |
| Eye problem | 56d | 49d | Worthless | 35 | 32 |
| Skin problem | 56e | 49e | Nervous | 45 | 40 |
| Stomaches | 56f | 49f | Lacks self-confidence | 47 | 42 |
| Vomiting | 56g | 49g | Fearful or anxious | 50 | 45 |
| Heart pounds | 56h | 49h | Feels guilty | 52 | 23 |
| Numbness | 56i | 49i | Self-conscious | 71 | 62 |
| **Thought Problems** |  |  | Sad | 103 | 93 |
| Hears things | 40 | 36 | Worries | 112 | 100 |
| Twitches | 46 | 41 | **Other Problems** |  |  |
| Repeats acts | 66 | 57 | Bored | 83 | 115 |
| Sees things | 70 | 61 | Shy | 75 | 66 |
| Strange behavior | 84 | 74 |  |  |  |
| Strange ideas | 85 | 75 |  |  |  |

**Table S1b.** Mapping of Older Adult Self Report (OASR) and Adult Self Report (ASR) common items that are assigned to different subscales.

| Question | ASR |  | OASR |  | Exact wording of question when not exact match |  |
| --- | --- | --- | --- | --- | --- | --- |
|  | Item # | Subscale | Item # | Subscale | ASR | OASR |
| Can't get mind off | 9 | Thought problems | 8 | Anxious/Depressed |  |  |
| Can't sit still | 10 | Other problems | 9 | Anxious/Depressed |  |  |
| Fidgety | 115 | Other problems | 34 | Anxious/Depressed |  |  |
| Dependent | 11 | Attention problems | 10 | Functional Impairment |  |  |
| Clumsy | 62 | Other problems | 55 | Functional Impairment |  |  |
| Sleep problem | 100 | Somatic complaints | 90 | Worries |  |  |
| Lacks energy | 102 | Attention problems | 92 | Functional Impairment |  |  |
| Argues | 3 | Aggressive behavior | 2 | Irritable/Disinhibited |  |  |
| Mean to others | 16 | Aggressive behavior | 15 | Irritable/Disinhibited |  |  |
| Demands attention | 19 | Intrusive | 18 | Irritable/Disinhibited |  |  |
| Not get along | 25 | Withdrawn | 22 | Irritable/Disinhibited |  |  |
| Badly with family | 28 | Aggressive behavior | 25 | Irritable/Disinhibited |  |  |
| Impulsive | 41 | Rule-breaking | 37 | Irritable/Disinhibited |  |  |
| Not liked | 48 | Withdrawn | 43 | Irritable/Disinhibited |  |  |
| Screams | 68 | Aggressive behavior | 59 | Irritable/Disinhibited |  |  |
| Shows off | 74 | Intrusive | 65 | Irritable/Disinhibited |  |  |
| Irresponsible | 76 | Rule-breaking | 67 | Irritable/Disinhibited |  |  |
| Stubborn | 86 | Aggressive behavior | 76 | Irritable/Disinhibited |  |  |
| Talks too much | 93 | Intrusive | 83 | Irritable/Disinhibited |  |  |
| Temper | 95 | Aggressive behavior | 85 | Irritable/Disinhibited | I have a hot temper | I lose my temper |
| Thinks about sex | 96 | Other problems | 86 | Irritable/Disinhibited |  |  |
| Loud | 104 | Intrusive | 94 | Irritable/Disinhibited |  |  |
| Can't concentrate | 8 | Attention problems | 7 | Memory/Cognition | I have trouble concentrating or paying attention for long | I have trouble concentrating or paying attention |
| Confused | 13 | Anxious/Depressed | 12 | Memory/Cognition |  |  |
| Can't decide | 78 | Attention problems | 69 | Memory/Cognition |  |  |
| Harms self | 18 | Thought problems | 17 | Other problems |  |  |
| Feels tired | 54 | Somatic complaints | 48 | Other problems |  |  |
| Attacks | 57 | Aggressive behavior | 50 | Other problems |  |  |
| Enjoys little | 60 | Withdrawn | 53 | Other problems |  |  |
| Steals | 82 | Rule-breaking | 73 | Other problems |  |  |
| Gets drunk | 90 | Rule-breaking | 80 | Other problems |  |  |
| Thinks suicide | 91 | Anxious/Depressed | 81 | Other problems |  |  |
| Trouble with the law | 92 | Rule-breaking | 82 | Other problems |  |  |
| Threatens | 97 | Aggressive behavior | 87 | Other problems |  |  |
| Can't succeed | 107 | Anxious/Depressed | 97 | Other problems | I feel that I can't succeed | I feel that I can't succeed at things |
| Jealous | 27 | Other problems | 24 | Thought problems |  |  |
| Badly with neighbors | 38 | Other problems | 27 | Thought problems |  |  |
| Out to get me | 34 | Anxious/Depressed | 31 | Thought problems |  |  |
| Rather be alone | 42 | Withdrawn | 38 | Thought problems |  |  |
| No friends | 67 | Withdrawn | 58 | Thought problems |  |  |
| Secretive | 69 | Withdrawn | 60 | Thought problems |  |  |
| Mood changes | 87 | Aggressive behavior | 77 | Thought problems |  |  |
| Withdrawn | 111 | Withdrawn | 99 | Thought problems |  |  |
| Fears | 29 | Other problems | 26 | Anxious/Depressed | I am afraid of certain animals, situations, or places | I am afraid of certain situations or places |
| Worries family | 72 | Other problems | 72 | Worries |  |  |
| Poor work | 61 | Attention problems | 54 | Functional Impairment | My work performance is poor | My performance at tasks is poor |
| Fails to finish | 59 | Attention problems | 52 | Memory/Cognition | I fail to finish things I should do | I have trouble finishing things I should do |
| Unloved | 33 | Anxious/Depressed | 30 | Thought problems | I feel that no one loves me | I feel that no one cares about me |
| Won't talk | 65 | Withdrawn | 56 | Other problems | I refuse to talk | I avoid talking |

**Table S2a.** Questions unique to Adult Self Report (ASR).

| Item # | Question | Item # | Question | Item # | Question |
| --- | --- | --- | --- | --- | --- |
| **Anxious/Depressed** |  | **Withdrawn** |  | **Aggressive behavior** |  |
| 113 | Worries opp. Sex | 30 | Poor opp. Sex | 5 | Blames |
| **Other problems** |  | **Attention problems** |  | 37 | Fights |
| 10 | Can't sit still | 1 | Too forgetful | 55 | Elation-depression |
| 21 | Damages others' things | 17 | Daydreams | 81 | Behavior changes |
| 24 | Doesn't eat well | 53 | Can't plan | 116 | Upset |
| 32 | Must be perfect | 64 | Can't prioritize | 118 | Impatient |
| 44 | Overwhelmed | 101 | Avoids work | **Rule-breaking behavior** |  |
| 58 | Picks skin | 105 | Disorganized | 6 | Uses drugs |
| 75 | Shy | 108 | Loses things | 20 | Damages own |
| 77 | Sleeps more | 119 | Poor at details | 23 | Breaks rules |
| 79 | Speech problems | 121 | Late | 26 | Lacks guilt |
| 89 | Rushes into things | **Intrusive** |  | 39 | Bad companions |
| 99 | Dislikes staying | 7 | Brags | 43 | Lies, cheats |
| 110 | Be opposite sex | 94 | Teases | 114 | Debts |
| 120 | Drives fast | **Thought problems** |  | 117 | Can't manage |
|  |  | 36 | Gets hurt | 122 | Can't keep job |
|  |  | 63 | Prefers older |  |  |

**Table S2b.** Questions unique to Older Adult Self Report (OASR).

| Item # | Question | Item # | Question |
| --- | --- | --- | --- |
| **Anxious/Depressed** |  | **Functional Impairment** |  |
| 14 | Concerned getting old | 3 | Getting things done |
| 21 | Worries future | 16 | Sits around |
| 28 | Fears doing bad | 29 | Preparing meals |
| 91 | Thinks about past | 68 | Sleeps more |
| 109 | Concerned health | 104 | Trouble dressing |
| **Worries** |  | 106 | Trouble bathing |
| 51 | Worries appearance | 111 | Soiling accidents |
| 89 | Concerned neatness | **Irritable/Disinhibited** |  |
| 101 | Wake up early | 19 | Damages things |
| 102 | Worries health | 35 | Wants own way |
| 117 | Gets too tired | 39 | Does things others don't like |
| 121 | Feels burdensome | 79 | Suspicious |
| **Somatic complaints** |  | 84 | Irritates people |
| 5 | Too much meds | **Other problems** |  |
| 33 | Feels sick | 49k | Other physical problems |
| 103 | Nightmares | 63 | Being punished |
| **Memory/Cognition problems** |  | 73 | Steals |
| 20 | Forget names | 105 | Doesn't like to phone |
| 70 | Can't talk |  |  |
| 110 | Can't remember |  |  |
| 114 | Forgets if not written |  |  |
| 122 | Worries memory |  |  |

**Table S3.** Baseline descriptors for each analyte from the blood sample collection. HbA1c = Hemoglobin A1C; HDL = High-density lipoprotein; LDL= Low-density lipoprotein; CRP = C-reactive protein; ESR = Erythrocyte sedimentation rate.

| Lab | n | Median | IQR | [5, 95%ile] |
| --- | --- | --- | --- | --- |
| Glucose | 279 | 92 | 15 | [77.9, 122.3] |
| HbA1c | 274 | 5.4 | 0.5 | [4.9, 6.535] |
| Cholesterol | 279 | 208 | 49.5 | [150, 274] |
| Triglyceride | 279 | 93 | 59 | [45, 210.3] |
| HDL | 275 | 63 | 27.5 | [38.7, 97] |
| LDL | 273 | 121 | 51 | [70.6, 179.4] |
| Albumin | 279 | 4.5 | 0.3 | [4, 4.81] |
| CRP | 276 | 0.1 | 0.2 | [0.1, 0.8] |
| Ferritin | 275 | 64 | 91 | [9, 217.6] |
| ESR | 278 | 8 | 12 | [2, 45.45] |
| Hemoglobin | 278 | 13.6 | 1.6 | [11.49, 15.82] |
| Hematocrit | 278 | 40.6 | 4.675 | [34.89, 46.82] |

**Table S4.** Intraclass Correlation Coefficients (ICCs) for phenotypic measures. CI = Confidence Intervals; See Figure 8 for definitions of variable abbreviations. Only baseline data are included and no filtering was done.

| Variable | n | ICC agreement | agreement 95% CI | ICC consistency | consistency 95% CI | cohen's d slope | t | p |
| --- | --- | --- | --- | --- | --- | --- | --- | --- |
| WASI BD | 198 | 0.90 | [0.87, 0.92] | 0.90 | [0.88, 0.92] | 0.22 | 3.05 | 0.0026 |
| WASI VOC | 198 | 0.79 | [0.75, 0.83] | 0.83 | [0.79, 0.86] | 0.71 | 9.94 | <.0001 |
| WASI MR | 198 | 0.77 | [0.72, 0.81] | 0.77 | [0.72, 0.82] | 0.21 | 2.98 | 0.0033 |
| WASI SIM | 197 | 0.67 | [0.61, 0.73] | 0.71 | [0.65, 0.76] | 0.53 | 7.48 | <.0001 |
| MoCA | 115 | 0.63 | [0.54, 0.72] | 0.64 | [0.55, 0.73] | 0.33 | 3.51 | 0.0006 |
| RAVLT | 177 | 0.72 | [0.66, 0.77] | 0.76 | [0.71, 0.81] | 0.70 | 9.25 | <.0001 |
| DKEFS CF | 179 | 0.72 | [0.66, 0.78] | 0.73 | [0.67, 0.78] | 0.19 | 2.59 | 0.0103 |
| DKEFS TMT | 197 | 0.74 | [0.69, 0.79] | 0.75 | [0.69, 0.80] | -0.20 | -2.78 | 0.006 |
| ANT Alert | 192 | 0.38 | [0.30, 0.47] | 0.39 | [0.30, 0.48] | 0.27 | 3.74 | 0.0002 |
| ANT Orienting | 191 | 0.16 | [0.08, 0.26] | 0.17 | [0.08, 0.26] | 0.20 | 2.70 | 0.0076 |
| ANT Conflict | 190 | 0.48 | [0.40, 0.56] | 0.5 | [0.42, 0.58] | -0.38 | -5.21 | <.0001 |
| ANT Grd Mean | 190 | 0.66 | [0.59, 0.72] | 0.67 | [0.61, 0.73] | -0.34 | -4.70 | <.0001 |
| MPraxis | 170 | 0.72 | [0.66, 0.78] | 0.72 | [0.66, 0.78] | 0.18 | 2.35 | 0.0197 |
| CET | 170 | 0.74 | [0.68, 0.80] | 0.74 | [0.68, 0.79] | 0.05 | 0.65 | 0.5155 |
| 2Back | 170 | 0.47 | [0.38, 0.56] | 0.47 | [0.38, 0.56] | 0.14 | 1.80 | 0.073 |
| CPT | 171 | 0.49 | [0.40, 0.57] | 0.51 | [0.42, 0.59] | 0.42 | 5.49 | <.0001 |
| BMI | 204 | 0.95 | [0.93, 0.96] | 0.95 | [0.93, 0.96] | -0.06 | -0.81 | 0.4186 |
| HbA1c | 132 | 0.74 | [0.67, 0.80] | 0.74 | [0.67, 0.80] | 0.39 | 4.46 | <.0001 |
| Cholesterol | 137 | 0.64 | [0.56, 0.71] | 0.65 | [0.57, 0.72] | -0.25 | -2.89 | 0.0045 |
| Pegboard | 200 | 0.68 | [0.62, 0.74] | 0.69 | [0.63, 0.75] | -0.26 | -3.70 | 0.0003 |
| Grip | 196 | 0.78 | [0.73, 0.82] | 0.78 | [0.73, 0.82] | -0.07 | -1.01 | 0.3129 |
| WWT WalkTime | 154 | 0.46 | [0.36, 0.55] | 0.47 | [0.38, 0.57] | 0.28 | 3.41 | 0.0008 |
| VO2max | 174 | 0.83 | [0.79, 0.87] | 0.83 | [0.79, 0.87] | -0.04 | -0.58 | 0.5632 |
| BDI | 199 | 0.75 | [0.69, 0.79] | 0.75 | [0.69, 0.79] | -0.08 | -1.07 | 0.2858 |
| STAI | 203 | 0.81 | [0.76, 0.84] | 0.81 | [0.76, 0.84] | -0.09 | -1.35 | 0.1787 |
| PSQI | 133 | 0.64 | [0.55, 0.71] | 0.64 | [0.55, 0.72] | -0.01 | -0.16 | 0.8712 |
| CogFailureQuest | 157 | 0.79 | [0.74, 0.84] | 0.81 | [0.76, 0.85] | -0.37 | -4.68 | <.0001 |
| SocialNetwork | 155 | 0.64 | [0.56, 0.71] | 0.64 | [0.57, 0.71] | -0.06 | -0.79 | 0.4326 |

## Supplementary Methods

### Full method description for diffusion MRI processing in QSIPrep

Preprocessing was performed using *QSIPrep* 0.23.1.dev0+g634483f.d20240830, which is based on *Nipype* 1.8.6 ^80^.

A T1w-reference map was computed after registration of 2 T1w images (after INU-correction) using antsRegistration [ANTs 2.4.3]. The anatomical reference image was reoriented into AC-PC alignment via a 6-DOF transform extracted from a full Affine registration to the MNI152NLin2009cAsym template. A full nonlinear registration to the template from AC-PC space was estimated via symmetric nonlinear registration (SyN) using antsRegistration. Brain extraction was performed on the T1w image using SynthStrip ^81^ and automated segmentation was performed using SynthSeg ^82^ from FreeSurfer version 7.3.1.

### Diffusion data preprocessing

Any images with a b-value less than 100 s/mm^2 were treated as a *b*=0 image. MP-PCA denoising as implemented in MRtrix3’s dwidenoise^83^ was applied with a auto-voxel window. When phase data were available, this was done on complex-valued data. After MP-PCA, Gibbs unringing was performed using MRtrix3’s mrdegibbs^84^. Following unringing, the mean intensity of the DWI series was adjusted so all the mean intensity of the b=0 images matched across each separate DWI scanning sequence. B1 field inhomogeneity was corrected using dwibiascorrect from MRtrix3 with the N4 algorithm^85^ after corrected images were resampled.

FSL (version None)’s eddy was used for head motion correction and Eddy current correction.^86^ Eddy was configured with a *q*-space smoothing factor of 10, a total of 5 iterations, and 1000 voxels used to estimate hyperparameters. A linear first level model and a linear second level model were used to characterize Eddy current-related spatial distortion. *q*-space coordinates were forcefully assigned to shells. Field offset was attempted to be separated from subject movement. Shells were aligned post-eddy. Eddy’s outlier replacement was run.^87^ Data were grouped by slice, only including values from slices determined to contain at least 250 intracerebral voxels. Groups deviating by more than 4 standard deviations from the prediction had their data replaced with imputed values. Final interpolation was performed using the jac method.

Several confounding time-series were calculated based on the preprocessed DWI: framewise displacement (FD) using the implementation in *Nipype* (following the definitions by^88^). The head-motion estimates calculated in the correction step were also placed within the corresponding confounds file. Slicewise cross correlation was also calculated. The DWI time-series were resampled to ACPC, generating a *preprocessed DWI run in ACPC space* with 1mm isotropic voxels.

Many internal operations of *QSIPrep* use *Nilearn* 0.10.1^89^; RRID:SCR_001362) and *Dipy*^90^. For more details of the pipeline, see <u>the section corresponding to workflows in *QSIPrep*’s</u> <u>documentation</u>.

Reconstruction was performed using *QSIRecon* 1.0.1.dev0+gf78c888.d20250114 (Cieslak et al. (2021)), which is based on *Nipype* 1.9.1 (Gorgolewski et al. (2011); Gorgolewski et al. (2018); RRID:SCR_002502).

### Anatomical data for DWI reconstruction

Brainmasks from antsBrainExtraction were used in all subsequent reconstruction steps. DSI Studio Reconstruction: Diffusion orientation distribution functions (ODFs) were reconstructed using generalized q-sampling imaging (GQI, Yeh, Wedeen, and Tseng (2010)) with a ratio of mean diffusion distance of 1.250000 in DSI Studio (version 94b9c79).

### DSI Studio Automatic Tractography

Automatic Tractography was run in DSI Studio (version 94b9c79) and bundle shape statistics were calculated (Yeh 2020). AutoTrack attempted to reconstruct the following bundles:

- Association_ArcuateFasciculus
- Association_Cingulum
- Association_ExtremeCapsule
- Association_FrontalAslantTract
- Association_HippocampusAlveus
- Association_InferiorFrontoOccipitalFasciculus
- Association_InferiorLongitudinalFasciculus
- Association_MiddleLongitudinalFasciculus
- Association_ParietalAslantTract
- Association_SuperiorLongitudinalFasciculus
- Association_UncinateFasciculus
- Association_VerticalOccipitalFasciculus
- Commissure_AnteriorCommissure
- Commissure_CorpusCallosum
- ProjectionBasalGanglia_AcousticRadiation
- ProjectionBasalGanglia_AnsaLenticularis
- ProjectionBasalGanglia_AnsaSubthalamica
- ProjectionBasalGanglia_CorticostriatalTract
- ProjectionBasalGanglia_FasciculusLenticularis
- ProjectionBasalGanglia_FasciculusSubthalamicus
- ProjectionBasalGanglia_Fornix
- ProjectionBasalGanglia_OpticRadiation
- ProjectionBasalGanglia_ThalamicRadiation
- ProjectionBrainstem_CorticobulbarTract
- ProjectionBrainstem_CorticopontineTract
- ProjectionBrainstem_CorticospinalTract
- ProjectionBrainstem_DentatorubrothalamicTract-lr
- ProjectionBrainstem_DentatorubrothalamicTract-rl
- ProjectionBrainstem_MedialForebrainBundle
- ProjectionBrainstem_MedialLemniscus
- ProjectionBrainstem_NonDecussatingDentatorubrothalamicTract
- ProjectionBrainstem_ReticularTract

Many internal operations of *QSIRecon* use *Nilearn* 0.10.1 (Abraham et al. 2014, RRID:SCR_001362) and *Dipy* 1.8.0(Garyfallidis et al. 2014). For more details of the pipeline, see <u>the section corresponding to workflows in *QSIRecon*’s documentation</u>.

